# ToolCAP: Novel Tools to improve management of paediatric Community-Acquired Pneumonia - a randomized controlled trial, Statistical Analysis Plan

**DOI:** 10.1101/2025.06.30.25329845

**Authors:** Silvia Cicconi, Jacques du Toit, Moniek Bresser, Fatimah Dhalla, Papa Moctar Faye, Luxi Lal, Helene Langet, Karim Manji, Andre Moser, Mama Awe Ndao, Megan Palmer, Jean Augustin Diegane Tine, Niel van Hoving, Kristina Keitel, Tracy Glass

## Abstract

The ToolCAP cohort study is a prospective, observational, multi-site platform study designed to collect harmonized, high-quality clinical, imaging, and biological data on children with IMCI-defined pneumonia in low- and middle-income countries (LMICs). The primary objective is to inform the development and validation of diagnostic and prognostic tools, including lung ultrasound (LUS), point-of-care biomarkers, and AI-based models, to improve pneumonia diagnosis, management, and antimicrobial stewardship. This statistical analysis plan (SAP) outlines the analytic strategy for describing the study population, assessing the performance of candidate diagnostic tools, and enabling data sharing in support of secondary research questions and AI model development.

Children under 12 years presenting with suspected pneumonia are enrolled within 24 hours of presentation and undergo clinical assessment, digital auscultation, LUS, and optional biological sampling. Follow-up occurs on Day 8 and Day 29 to assess outcomes including recovery, treatment response, and complications. The SAP details variable definitions, data management strategies, and pre-specified analyses, including descriptive summaries, sensitivity and specificity of diagnostic tools against clinical reference standards, and exploratory subgroup analyses.

## 1 Administrative information

### 1.1 Document scope

This document provides a detailed description of the methodologies that will be followed, as closely as possible, when analyzing and reporting results from ToolCAP study. The planned analysis detailed in this document is in compliance with that briefly specified in the ToolCAP protocol, except if otherwise detailed.

The purpose of this Statistical Analysis Plan (SAP) is to:

- Ensure that the analysis is appropriate for the aims of the study, reflects good statistical practice in general, and minimizes bias.
- Ensure that the analyses performed are consistent with the conditions of the protocol.
- Explain in detail how the data will be handled, endpoints and covariates derived and analyzed to ensure transparency and reproducibility, including enabling others to perform the actual analysis in the event of sickness or other absence.
- Protect the project by helping it keep to timelines and within scope.
- Allow analysis reproducibility once study data are shared in the public domain after study closure and final analysis is completed.

Additional exploratory or auxiliary analyses of data not specified in the protocol or this document are permitted but fall outside the scope of this SAP. Where analyses are presented which are not included in the SAP, these will be clearly indicated as such along with a justification as to their inclusion.

The analysis documents will be made available if required by journals or referees when the main papers are submitted for publication. Additional analyses suggested by reviewers or editors will, if considered appropriate, be performed in accordance with the analysis plan, but if reported, the source of such post-hoc analyses shall be declared.

This document is written following the guidelines outlined in Gamble et al [1], the CONSORT statement [2] and ICH-e9 guidelines [3].

### 1.2 Document history

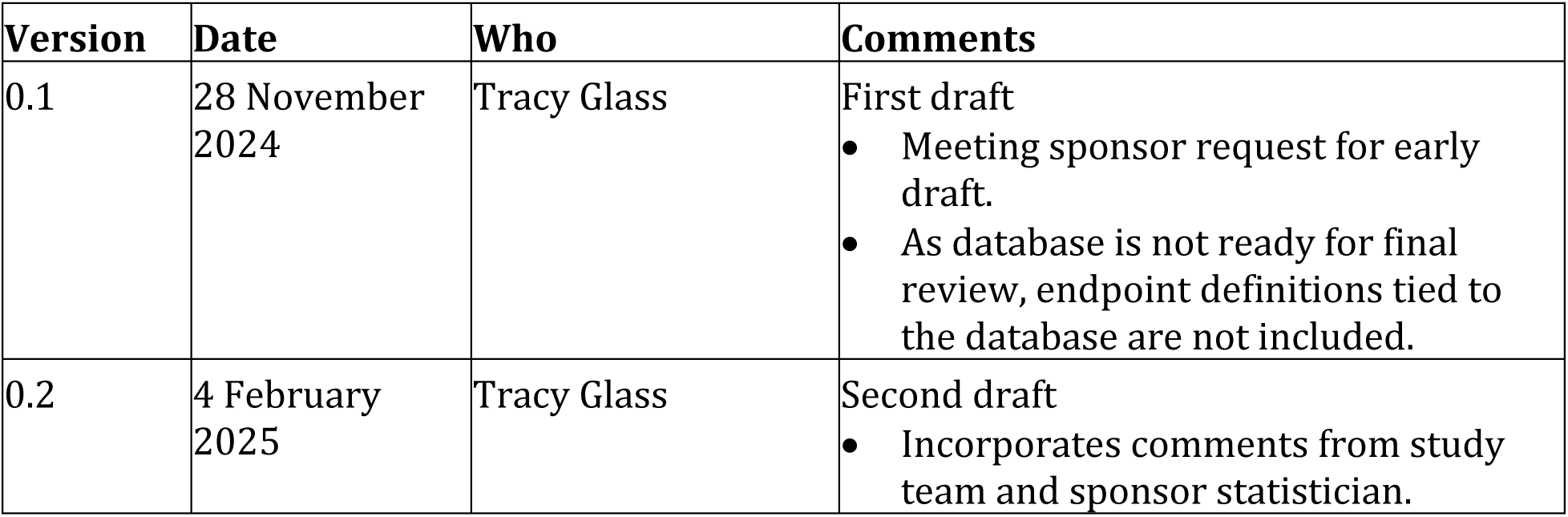
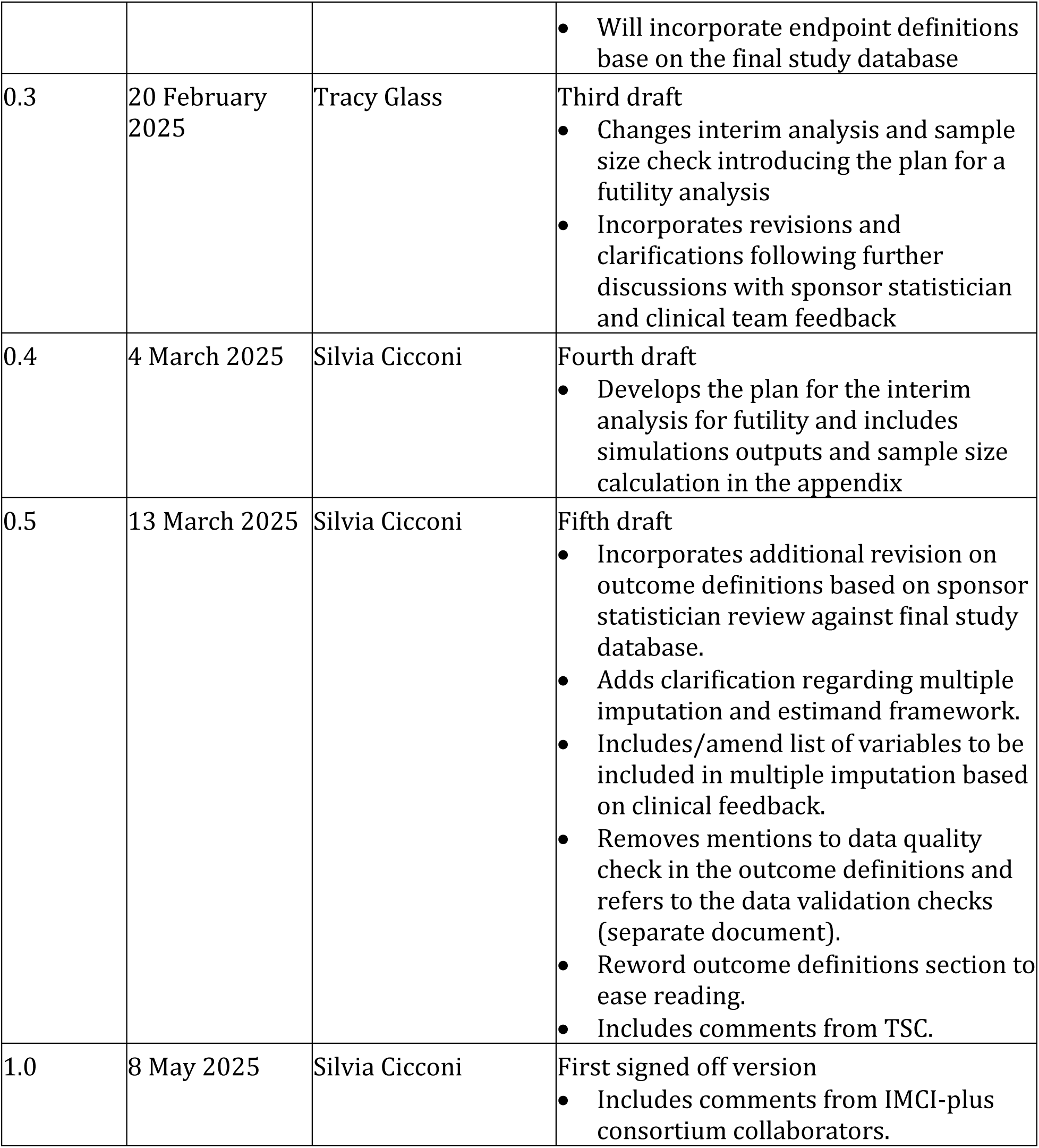

#### Roles and responsibilities

| Name | Affiliation | Role |
| --- | --- | --- |
| Tracy Glass | Swiss TPH, Switzerland | Senior trial statistician |
| Kristina Keitel | University of Bern, Switzerland | Sponsor Representative & Overall Coordinating Investigator |
| Jacques du Toit | MRC/Wits Rural Public Health and Health Transitions Research Unit (Agincourt), South Africa | Coordinating Investigator Africa & Principal Investigator. Wits University, South Africa |
| Stephen Tollman | MRC/Wits Rural Public Health and Health Transitions Research Unit (Agincourt), South Africa | Principal Investigator.<br>Wits University, South Africa |
| Karim Manji | Muhimbili University of Health and Allied Sciences, Tanzania | Principal Investigator, MUHAS, Tanzania |
| Nayha Salim | Muhimbili University of Health and Allied Sciences, Tanzania | Principal Investigator, MUHAS, Tanzania |
| Megan Palmer | Desmond Tutu TB Centre, South Africa | Principal Investigator, Stellenbosch University, South Africa |
| Niel van Hoving | Stellenbosch University, South Africa | Principal Investigator, Stellenbosch University, South Africa |
| Papa Moctar Faye | Université Cheikh Anta Diop Dakar, Sénégal | Principal Investigator, Université Cheikh Anta Diop de Dakar (UCAD), Senegal |
| Moniek Bresser | Swiss TPH, Switzerland | Data manager RCT |
| Silvia Cicconi | Swiss TPH, Switzerland | Trial statistician |
| Daniel Ohene-Kwofie | MRC/Wits Rural Public Health and Health Transitions Research Unit (Agincourt), South Africa | Data manager lead |
| Yasir Nisar | World Health Organization | Trial Steering Committee, Chairman |
| Jonathan Levin | WITS University | Trial Steering Committee, External member |

##### Signatures

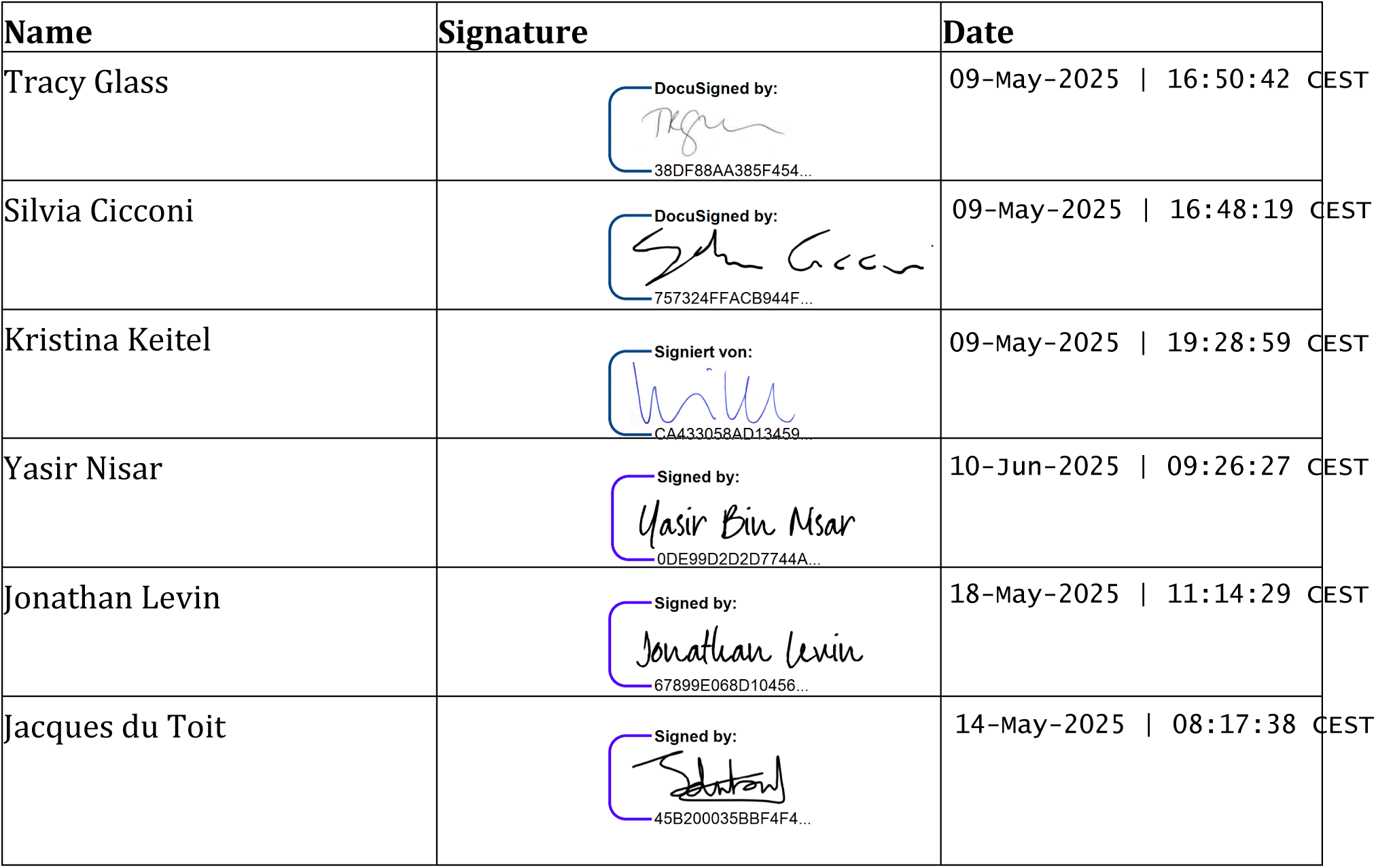

#### Data management and sharing

Detailed information is provided in the Data Management Plan (DMP). Briefly, data are captured by study staff directly into an electronic REDCap database, set up by MRC/Wits-Agincourt Research Unit as designated by the Sponsor Representative and hosted at Wits University in South Africa (a copy only for Tanzania data, which are primary hosted at MUHAS). Data entered into the eCRF is automatically validated for completeness and discrepancies. Additional pre-defined data quality checks are detailed in the Data Review Log. Queries raised as a result of these checks will be resolved by the site teams.

The study team of a local site will only have access to the data of the participants and to the data entered in this center. The study team of a local site will access the data through a tablet computer or a computer. Access to the secured platform will be password-protected.

De-identified experimental array data will be preserved in CSV format and accompanying DDI files and files in line with UK Data Service recommendations. This data will be stored in academic data repositories such as the Agincourt data repository or Zenodo, which is an open-access repository operated by CERN and part of the European OpenAIRE project (see Section 4.3 and 4.4 of the DMP).

## 2. Introduction

### 2.1 Background and rationale

In 2019, lower respiratory tract infections (LRTI) caused 2.6 million deaths worldwide, making them the fourth leading cause of death overall. LRTI (including pneumonia) are the most common reason for sick children to present for acute outpatient care. Current best practice pneumonia management guidelines advocate for all cases to receive a course of antibiotics, based on the World Health Organization’s (WHO) Integrated Management of Childhood Infections (IMCI) framework. The framework relies on presumptive antibiotic treatment based on clinical signs alone (cough and/or difficulty breathing, respiratory rate, and lower chest wall costal indrawing) despite these clinical features inadequately distinguishing bacterial causes of pneumonia from the vastly more common, self-limiting viral infections.

Improving IMCI diagnostic criteria to better identify children with pneumonia that would benefit from antibiotic therapy is therefore a WHO research priority, along with a growing interest in the potential of Lung Ultrasound (LUS) to address this. LUS has now become increasingly affordable in low-resource-settings thanks to recent innovations in its “ultrasound-on-a-chip” design; it could be integrated into the standard clinical exam without incurring extra costs, time, radiation, or specialist consultation. This, together with increasing evidence showing its ability to effectively detect lung consolidation in pneumonia, have made it an attractive tool for frontline clinicians.

We therefore propose that the integration of lung ultrasound (LUS) (including standardised interpretation) into the IMCI-based management framework of clinical pneumonia will better identify those who would benefit from antibiotic treatment, which will lead to a reduction in antibiotic overuse without compromising health outcomes.

Please refer to the section Background and rationale of the Protocol for more information.

### 2.2 Objectives

The overall goal of the ToolCAP study is to reduce antibiotic prescriptions in children with WHO IMCI clinical pneumonia attending primary care facilities, while having no detrimental effect on clinical failure in these children. The study will compare the introduction of LUS (“intervention”) for the assessment of treatment to the current standard of care (“control”) in children aged 60 days - 12 years. We hypothesize that the antibiotic prescription rate will be lower in the intervention arm and the clinical failure rate by day 8 will be non-inferior between the intervention and control arms.

## 3. Study methods

### 3.1 Study design

#### 3.1.1 Overview

This is a prospective, open label, individually randomized controlled trial (RCT) with two arms (intervention and control) investigating Lung Ultrasound (LUS) as a novel tool for the management of WHO clinical pneumonia in outpatient and emergency room settings (protocol to be published).

#### 3.1.2 Study Procedures

Participants will be consecutively enrolled and randomized in the ratio 1:1. All paediatric patients (60 days – 12 years) presenting to the ER or Outpatients Department (OPD) at participating study sites will be triaged as per routine care. Study staff will screen all presenting patients and patients who meet the inclusion criteria and provide informed consent will be enrolled consecutively, as specified in the protocol (to be published).

#### 3.1.3 Study arms

The two arms of the trial are in brief:

“SOC” arm:

- The control group will be managed by treating clinicians as per routine care. Routine care in South African, Tanzanian, and Senegalese District/Regional Hospitals are as per their respective standard treatment guidelines [4, 5] which are based on IMCI for outpatient visits for children up to 5 years of age. For older children these guidelines are based on national recommendations. The IMCI framework recommends antibiotic prescriptions for all cases.

“Intervention” arm:

- The intervention will integrate LUS into current management guidelines for pneumonia (national guidelines based on IMCI). Patients will receive LUS on the day of enrolment and clinicians will be provided guidance on interpretation of the findings and the implications on antibiotic prescribing (see Figure 1). Additional guidance regarding admission or site of care will not be provided and will be based on local/national guidelines based on IMCI as per routine care.

**Figure 1:**
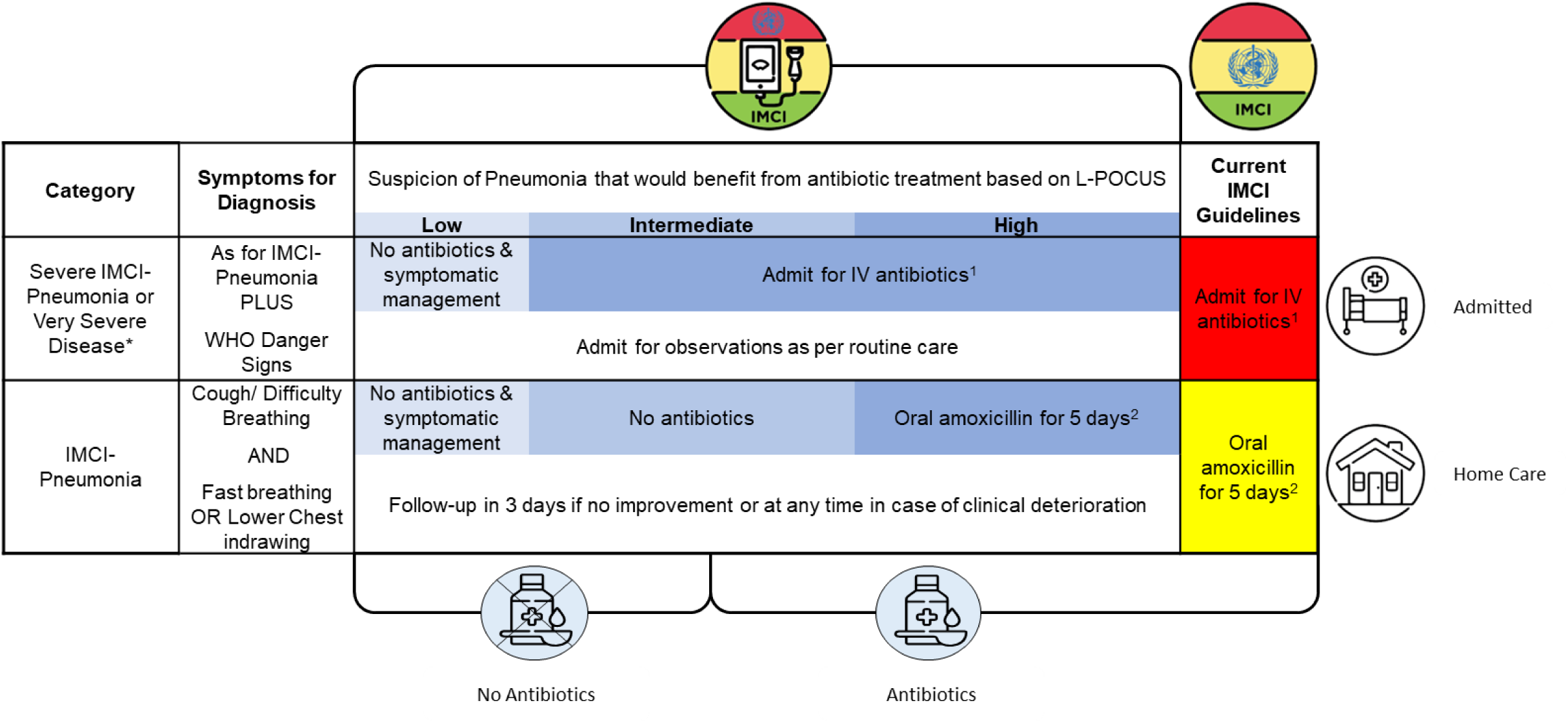
Summary of the IMCI-PLUS algorithm

Further information about procedures for the trial arms can be found in the protocol (to be published).

#### 3.1.3 Study outcomes

As per the protocol, the endpoints of the trial are as follows:

**Co-Primary Endpoints**

i) Proportion of participants prescribed antibiotic treatment in each study group on Day 1 (D1).
ii) Proportion of participants with clinical failure, defined as the development of any of the following criteria during the specified time periods after randomization.

a) Any time before or on Day 8 (D8):

- WHO IMCI danger sign (inability to drink/breastfeed, vomiting everything, convulsions with this illness, lethargy/unconscious)
- New or worsening severe respiratory distress (such as grunting, head nodding, severe chest indrawing)
- Secondary Hospitalization (defined as a hospitalization occurring after discharge from in-patient admission or outpatient visit) related to a deterioration of the presenting complaint on D1
- Change in level of care (e.g., admission to intensive care unit, transfer to higher level of care)
- Need for respiratory support (e.g., high flow nasal cannula, CPAP)
- Death due to any medical cause (i.e., except trauma)
b) At D8 outcome assessment:

- Report from the caregiver of non-resolution/worsening of illness

**Secondary endpoints:**

i) Evaluation of the effect of the intervention in children with clinical pneumonia on:

- The proportion of children prescribed an antibiotic by D8.
- The proportion of adverse drug reactions related to routine antibiotic treatment by D8 (i.e., Anaphylactic reaction, severe diarrhoea, or generalised severe rash).
- The proportion of patients cured (defined as caregiver reported recovery from illness) at D8 and Day 29 (D29).
- The proportion of patients admitted to hospital on D1 and by D8.
- The duration of the D1 inpatient admissions (up until D29).
- The duration of inpatient admissions starting between D1 and D8 (until D29)
- The proportion of patients undergoing a non-study related diagnostic test (including Chest Xray (CXR), blood tests, urine tests, microbiological assays) on D1, and for those admitted on D1, tests conducted up until D8.
- The proportion of deaths of any cause by D29.
- The proportion of participants with unscheduled health seeking events for any cause and/or hospital admission for any cause since D8 follow-up (at D29 follow-up).
ii) Proportion of different aetiological diagnoses (e.g., pulmonary tuberculosis, RSV bronchiolitis, confirmed bacterial pneumonia). – Pending protocol amendment to be moved to exploratory endpoints.
iii) In children enrolled in the TB substudy:

- The proportion of children with confirmed, unconfirmed or unlikely intrathoracic TB
- The proportion of children:

i. meeting exclusion criteria 1-9 and not enrolled in the RCT OR
ii. enrolled in the RCT and classified as having clinical failure who ultimately had TB disease
- The ability of LUS (and mediastinal US, if done) to distinguish TB from non-TB disease
- The diagnostic accuracy of LUS compared with CXR and CAD within the context of TB Treatment Decision Algorithms
iv) Cost-effectiveness of integration of LUS into current IMCI-based management framework for pneumonia (separate analysis to be detailed in separate SAP).

**Exploratory endpoints (listed here but analysis detailed in a separate SAP):**

1) Evaluation of the effect of the intervention in children with clinical pneumonia on:

a. The number of days of antibiotic treatment received between D1 and D8
b. The proportion of children with a change in antibiotic treatment strategy by D8 (including class switching, switching of agent, extension of spectrum).
2) Proportion and type of abnormalities detected in LAusc and their retrospective predictive performance (compared to clinical signs and symptoms, CXR and host biomarkers as available) in the diagnosis and risk stratification of pneumonia.
3) Sensitivity, specificity, positive predictive value (PPV) and negative predictive value (NPV) of LUS in comparison to CXR (Retrospective analysis).
4) The ability of a clinical LUS severity score in stratifying risk in a cohort of patients with clinical pneumonia.
5) Improved risk stratification and management guidelines for clinical pneumonia through the inclusion of improved LUS feature thresholds and novel and personalized biomarkers.
6) Identification of the various definitions of pneumonia (clinical, IMCI, expert LUS, retrospective diagnosis) using a deep learning algorithm using LUS (with and without other clinical variables).
7) Stratification of disease severity using a deep learning algorithm using LUS (+/-other clinical variables)
8) Proportion and type of abnormalities detected on LUS on blinded retrospective expert LUS analysis in control group.
9) In children enrolled in the TB sub-study:

a. The role of novel biomarkers in the diagnosis of TB in children
b. The proportion with sole or concomitant extrapulmonary TB

### 3.2 Randomization

Electronic randomization envelopes are used in the ToolCAP study. A dedicated REDCap database, available online, will host the randomization envelopes and the randomization lists as programmed by the trial statistician and generated by an independent statistician. The randomization lists will be stored in an access-restricted folder, accessible only to the independent statistician and the Data Oversight Team. More details can be found in the DMP.

Enrollment and randomization should occur on the same study day. The date of randomization will be considered as Day 1.

Participants will be randomized 1:1 to intervention and control. Participants will be randomized using block randomization with varying block sizes (details on block sizes held by the trial statistician), stratified by site (za_wc1, za_mp1, za_mp2 for South Africa, tz_mh1, tz_mh2, tz_mh3, tz_mh4 for Tanzania, and sn_uc1 for Senegal) and age groups (<5, 5-<13). If the full date of birth is not known, the participant is required to enter a value for years and months of age, therefore the age will be estimated with sufficient detail for the stratification. If the years of age cannot be reliably estimated, the participant should not be enrolled into the trial.

Once participant ID (unique identifier assigned at screening), enrollment date and age category (<5, 5-<13) have been confirmed in the REDCap randomization database, the randomization envelope displays an Envelope ID and the randomization allocation. Both Envelope ID and the randomization allocation will be entered in the clinical database.

### 3.3 Blinding

Study site blinding is not feasible in this trial and therefore it will be an open-label study. Hospital clinicians will not be able to perform LUS or receive the LUS findings for the control group as the use of LUS is not standard practice for routine management in the site hospitals. Only study staff will be conducting LUS and clinicians will be requested not to use LUS during the study.

Study personnel assessing the endpoints during follow-up will be blinded to the allocation to the best extent possible. However, during the follow-up, the parent or guardian may mention information that informs the allocation. This could introduce bias into the endpoint assessment for clinical failure and training will be provided to personnel to minimize this potential bias.

The trial statistician will be unblinded.

### 3.4 Sample size

An extensive literature review was done to inform the parameters for the sample size estimation. This document is provided in the Appendix A.

With an assumed baseline proportion of clinical failure in the control group of 5% and using an acceptable clinical difference of 1.5 as a non-inferiority margin (therefore an absolute delta of 2.5%), the estimated sample size required across all sites would be 2730 to achieve a power of 85% with a one-sided type 1 error of 2.5%. This would equate to 1365 participants per group for the co-primary endpoints. Taking into consideration an estimated lost to follow-up of 5%, repeat visits of the same child within the enrolment period, as well as potential differences in clinical failure rates between sites, we aimed to enroll 3000 participants.

As children under 5 years of age are the group of greatest interest based on the current IMCI framework, we powered the study to have sufficient power to test for differences in this subgroup. Assuming that children aged <5 years makes up 90% of the children presenting for acute respiratory care among those <13 years and wanting to add to the research in those >5 years, we aim to include 500 children from 5-12 years of age. Therefore, we aim for a total sample size of 3500 participants, 1750 participants per group.

For the second co-primary outcome with a superiority comparison, we expect an antibiotic prescription rate of 80% in the control arm. With a sample size of 3500 and a two-sided type 1 error of 5%, we would have 100% power to detect a decrease in antibiotic prescription rates of 10% or more. We consider a reduction in antibiotic prescriptions of 15% or more on day 1 to be clinically relevant.

Overall, the study will have a total power of 85% to show non-inferiority in terms of clinical failure by Day 8 and superiority in terms of reduction of antibiotic prescriptions on Day 1.

The planned breakdown of participant enrolment per country is as follows: Tanzania – 2000 participants, Senegal – 500 participants, South Africa – 1000 participants.

It is not planned to replace participants found to be ineligible post-randomization as this number is expected to be small. However this will be assessed and potentially reconsidered at the time of the interim sample size check (see section 3.6).

Although the protocol allows for multiple re-enrollment of the same child (after 29 days or more from the previous enrollment date), the sample size calculation is based on participants, not on presentations. Children who are enrolled multiple times contributes once to the target sample size and only the first presentation will be included in the primary analysis.

### 3.5 Framework

The co-primary endpoint comparisons between the two arms is superior in terms of reduction of antibiotic prescriptions on Day 1 and non-inferior in terms of clinical failure by Day 8.

We hypothesize that the intervention will result in a significantly lower antibiotic rate (by at least 15%) compared to the expected rate of 80% in the control arm (see 3.4).

H_0_: There is no difference between the experimental arm and control arm with respect to antibiotic prescription on day 1.
H_1_: There is a difference between the experimental arm and control arm with respect to antibiotic prescription on day 1.

We hypothesize that the intervention is non-inferior to the control arm with respect to clinical failure at Day 8 given an assumed control failure rate of 5% and a non-inferiority margin of 2.5% (see 3.4).

H_0_: The difference between the experimental arm and control arm with respect to clinical failure by day 8 is greater than the upper non-inferiority margin.
H_1_: The difference between the experimental arm and control arm with respect to clinical failure by day 8 is less than or equal to the upper non-inferiority margin.

### 3.6 Interim analyses

An independent data monitoring committee (IDMC) was set up to protect the safety of study participants, to assist and advise the ToolCAP trial steering committee (TSC) in order to protect the integrity, validity and credibility of the trial. Further details on the composition and role of the IDMC are provided in the IDMC charter.

The IDMC and TSC will be provided with regular updates by the data manager via email that include overall screening and recruitment by site, protocol adherence (inclusion of ineligible participants, if found), intervention adherence, baseline (gender, age group, and symptoms at enrollment), and follow-up (percent reached for Day 8 and 28 phone calls) summaries. The information are pooled over both intervention arms. In addition, the trial statistician will provide the IDMC with an open report (containing pooled data) and a closed report (containing data by arms) for the IDMC meetings.

Two IDMC meetings are foreseen:

- The first scheduled IDMC meeting will take place after the first 200 participants are recruited, corresponding to approximately 6-8 weeks after trial start. The open report for this meeting will contain the pooled information in the regular report update (see above), plus safety listings and summaries. A closed report with data summarized by arm will be provided to the IDMC members by the trial statistician for discussion during the closed session. In the closed report, the arms will be labelled as “A” and “B” and unblinded during the closed meeting at the discretion of the IDMC.
- The second scheduled IDMC meeting is planned after 50% of participants (1750) have been recruited. The content of the open and closed report for this meeting will be the same as for the first IDMC meeting (see point above) but also include:

◦ Expected and actual recruitment rates: to assess if we are on track to recruit the planned sample size in the expected time frame (and considering the outcome of the previous point). If recruitment is slower than expected (see section 5.3), the IDMC may recommend to consider adding sites to the trial.
◦ Eligibility and protocol deviations: to assess if there are procedural or unexpected reasons why individuals are only being identified as ineligible after randomization. If this is occurring, we will discuss with the IDMC whether these individuals should be excluded and replaced or if they will be kept and included in FAS (as planned). Additional information to assess sample size assumptions and conditional power (more details below).
◦ Information to assess sample size assumptions and conditional power (more details below).

The sample size re-estimation is based on a blinded re-estimation approach [22–23] meaning that both co-primary outcomes will be summarized only pooled and not by arm.

The sample size re-estimation is driven by clinical failure rate:

- If the observed clinical failure rate is higher than our assumed value, no sample size calculation will be done as no reduction in sample size is foreseen.
- If the observed clinical failure rate is less than the assumed clinical failure rate (5%), an increase in sample size would be needed. Note that the maximum feasible sample size increase was a priori limited to 5000 children. We will recalculate the sample size using the pooled estimates from the interim analysis [22], adapting the absolute NI margin while keeping the relative NI margin of 1.5 constant.

At the interim stage (50% of the planned children) we assess the conditional power to assess an early stopping because of futility, given the interim data accumulated up to the time of the interim analysis. In case of an increased re-estimated sample size, the updated sample size (or the defined maximum of 5,000 children) will be used for the conditional power. Otherwise, the initially planned sample size will be used. We assumed for future patients the initial sample assumptions (clinical failure rate=5%, antibiotics rate 80% in control group and 65% in experimental group). Under these assumptions the trial will be recommended to stop because of futility if the conditional power is lower than 20%. The type-I error is not affected under futility stop, but the power decreases. We also performed simulation studies to assess the power under futility stopping (see Appendix B). However, this is a non-binding rule and should be used by the IDMC as a guidance in making recommendation to the Sponsor as to continuing or stopping the study.

We performed simulation studies (see Appendix B) to evaluate different interim analysis scenarios and assess the impact of a sample size re-estimation and the power under futility stopping. Due to the change of the absolute NI margin [24], the type I error was also assessed in simulation studies.

Any changes to the sample size or recruitment strategy will be documented in a protocol amendment and in the final study report.

### 3.7 Timing of analyses

For the IDMC meetings, data will be analyzed after all children recruited (200 for the first and 1750 for the second) reach Day 8 follow-up and after data cleaning processes. The data export will include all data collected on these children up to the date of the export. The IDMC meeting should be scheduled a minimum of two weeks after the database is exported in order to have sufficient time to create the report and allow the IDMC time to review the report.

The final analysis will take place once all participants have reached the window for Day 29 follow-up. Once the database is considered complete and cleaned, it will be locked and the final analysis will commence.

### 3.8 Timing of baseline and outcome assessments

Baseline is defined as the day of randomization (D1) and time will be measured from this point.

The primary outcome of antibiotic prescription on D1 will be based on activities occurring only on D1 although the information about this event can come from information obtained during follow-up (see 6.1.1).

The information for the primary outcome of clinical failure will come from data collected on D1 through the D8 window (≤day 12). Information collected through D29 window (<34) referencing events occurring between D1 and D8 can be utilized if this information was not collected prior to this. In the case conflicting information is provided at D29, the earlier information provided through D8 will be utilized (see 6.1.1).

Table 1 shows the nominal activity days of the outcomes and the permitted ranges for their data collection as defined in the protocol.

**Table 1.**
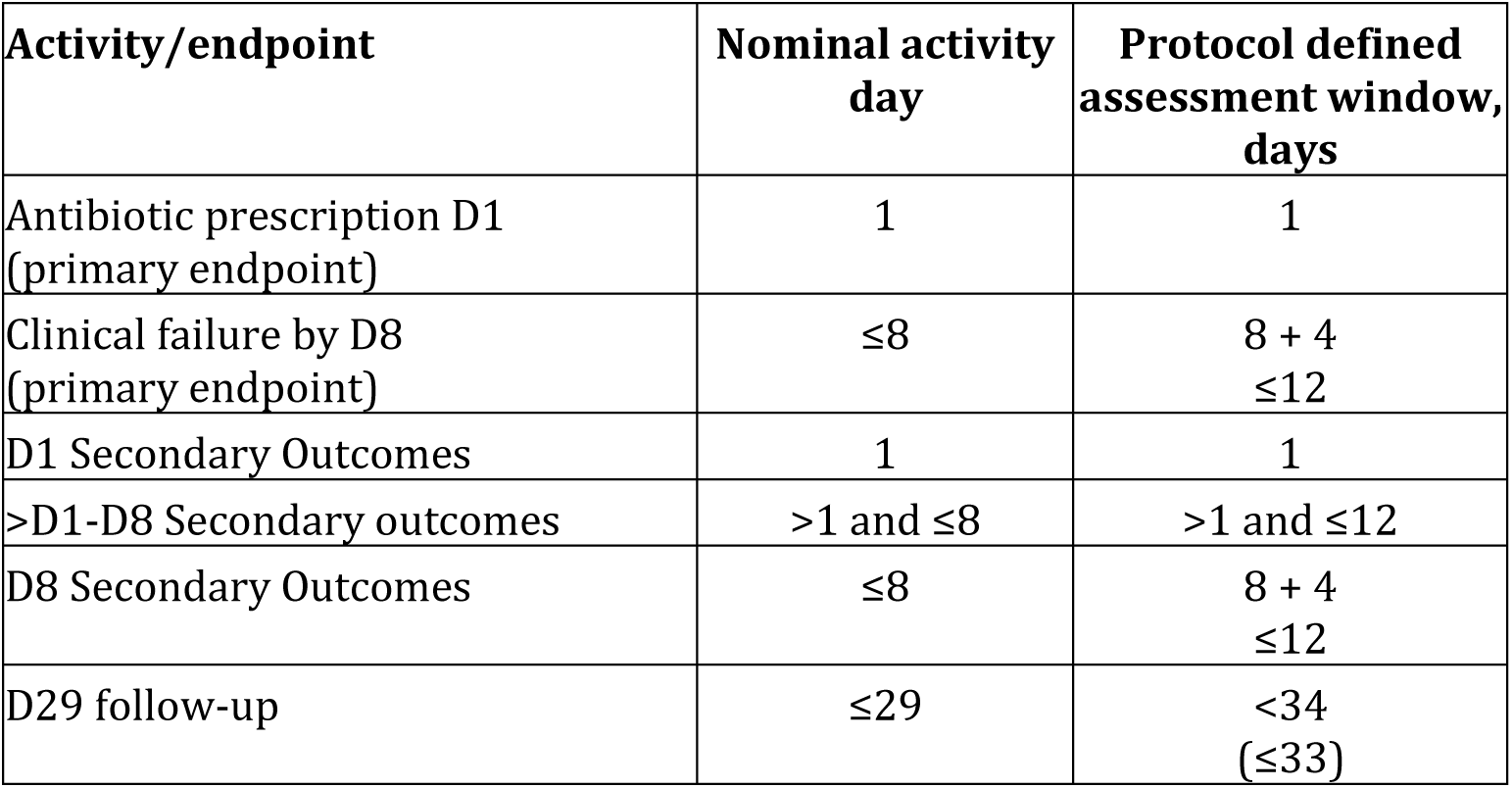
Nominal activities and permitted windows.

| Activity/endpoint | Nominal activity day | Protocol defined assessment window, days |
| --- | --- | --- |
| Antibiotic prescription D1 (primary endpoint) | 1 | 1 |
| Clinical failure by D8 (primary endpoint) | $\leq 8$ | 8 + 4<br>$\leq 12$ |
| D1 Secondary Outcomes | 1 | 1 |
| >D1-D8 Secondary outcomes | $>1$ and $\leq 8$ | $>1$ and $\leq 12$ |
| D8 Secondary Outcomes | $\leq 8$ | 8 + 4<br>$\leq 12$ |
| D29 follow-up | $\leq 29$ | $<34$<br>($\leq 33$) |

## 4. Statistical principles

### 4.1 Confidence intervals and p-values

Wald statistical tests will be one-sided for non-inferiority and two-sided for superiority hypotheses. Estimates will be presented with 95% two-sided Wald confidence intervals. P-values will be presented where appropriate for superiority comparisons only. No type-I adjustments will be made for as only a blinded sample size recalculation is implemented and stopping for futility does not inflate the type-I error. Interpretations of secondary outcomes will be based on the strength of evidence of effect size and consistency of results.

### 4.2 Adherence and protocol deviations

#### Adherence

We foresee several potential adherence issues in the study:

- Control arm: Clinicians did not follow the IMCI framework by not prescribing antibiotics to participants.
- Intervention arm: Clinicians did not follow the recommendation of the LUS for antibiotic prescription (lus_rept_abx on Lung Ultrasound report).

#### Protocol deviations

Protocol deviations for analysis considerations are defined as follows:

- Control arm: LUS was performed and the treating clinician had access to the results (pd_type = 4)
- Intervention arm: Results of the LUS were not provided to the clinician (indicated on LUS report) (pd_type = 4).
- Any participants found to be ineligible post-randomization. These cases will be monitored throughout the trial and procedures put in place to minimize their occurrence (pd_type=2) (see 3.6.1).

Any non-adherence to the protocol or guidelines as well as protocol deviations as reported by the trial team or detected at the time of data cleaning or analysis will be described by arm.

### 4.3 Analysis populations

#### Full Analysis Set (FAS)

The FAS should resemble as closely as possible the intent-to-treat principle and will include all randomized subjects in their assigned arms.

#### Per-protocol Set (PPS)

The PPS will exclude all participants with protocol deviations (see 4.2).

Primary analysis for the non-inferiority comparison of clinical failure rates will be performed on both the per-protocol (PP) and Full Analysis sets (FAS) [6]. Primary analysis of the superiority comparison of antibiotic prescribing rates will also be performed on both the FAS and PP set.

Our approaches for the primary outcome analysis are further detailed in the context of the estimands framework in Tables 2a-2d [7].

**Table 2a.** Estimand framework: First co-primary estimand for superiority comparison of antibiotic prescription using FAS population [1].

| Estimand attribute | Definition | Comments |
| --- | --- | --- |
| Study Population | Children aged 60 days to 12 years presenting for acute care to a study site with WHO IMCI clinical pneumonia. | Exclusion of children considered too ill to follow standard care (IMCI danger sign, jaundice, hypoxemia, requiring non-invasive ventilator support), presenting for a follow-up visit of a treated LRTI, having received antibiotics for more than 48 hours, or selected HIV-positive (<12 months old, uncontrolled or requiring admission) were excluded as written in the protocol. |
| Analysis Population | FAS | All baseline eligible participants. |
| Treatment conditions | Routine of care (“Control”) vs. use of LUS for treatment decision (“Intervention”) |  |
| Endpoint | Antibiotic prescribed on Day 1 |  |
| Summary measure | Risk differences. We will also report odds ratios and risk ratios. |  |
| Handling of intercurrent events (IE) [1] |  |  |
| Did not follow treatment assignment according to protocol | “Treatment policy” approach | The IE are defined as 1) not prescribing antibiotics in the “Control” arm and 2) not following the recommendations of the LUS in the “Intervention” arm. This IE is ignored and considered part of the intention to treat strategy |
| Death on D1 | “Principal stratum” approach | Those with the IE should likely not have been randomized as too ill to follow standard care (see comment under study population). Although it is possible they experienced the outcome prior to the IE, they are excluded as it is likely that information on the outcome would not be consistently recorded and introduce bias. |

[1] Intercurrent events are post-randomization events that affect either the interpretation or existence of the outcome data associated with the clinical question of interest (missing data or LTFU is not in itself an intercurrent event; see instead section 6.3 Missing data).

**Table 2b.** Estimand framework: First co-primary estimand for superiority comparison of antibiotic prescription using PP population [1].

| Estimand attribute | Definition | Comments |
| --- | --- | --- |
| Study Population | Children aged 60 days to 12 years presenting for acute care to a study site with WHO IMCI clinical pneumonia. | Exclusion of children considered too ill to follow standard care (IMCI danger sign, jaundice, hypoxemia, requiring non-invasive ventilator support), presenting for a follow-up visit of a treated LRTI, having received antibiotics for more than 48 hours, or selected HIV-positive (<12 months old, uncontrolled or requiring admission) were excluded as written in the protocol. |
| Analysis Population | PP | All baseline eligible participants excluding those with protocol deviations. |
| Treatment conditions | Routine of care (“Control”) vs. use of LUS for treatment decision (“Intervention”) |  |
| Endpoint | Antibiotic prescribed on Day 1 |  |
| Summary measure | Risk differences. We will also report odds ratios and risk ratios. |  |
| Handling of intercurrent events (IE) [1] |  |  |
| Death on D1 | “Principal stratum” approach | Those with the IE should likely not have been randomized as too ill to follow standard care (see comment under study population). Although it is possible they experienced the outcome prior to the IE, they are excluded as it is likely that information on the outcome would not be consistently recorded and introduce bias. |

[1] Intercurrent events are post-randomization events that affect either the interpretation or existence of the outcome data associated with the clinical question of interest (missing data or LTFU is not in itself an intercurrent event; see instead section 6.3 Missing data).

**Table 2c.**
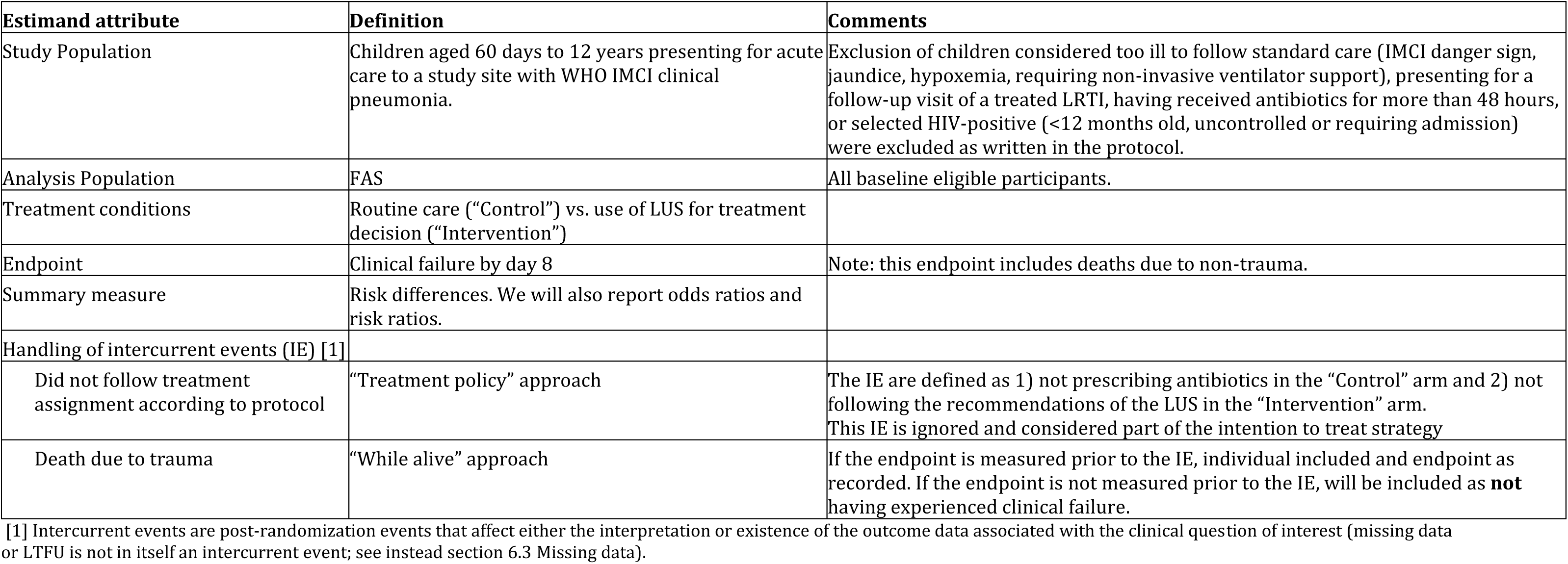
Estimand framework: second co-primary outcome of clinical failure using FAS population [1].

| Estimand attribute | Definition | Comments |
| --- | --- | --- |
| Study Population | Children aged 60 days to 12 years presenting for acute care to a study site with WHO IMCI clinical pneumonia. | Exclusion of children considered too ill to follow standard care (IMCI danger sign, jaundice, hypoxemia, requiring non-invasive ventilator support), presenting for a follow-up visit of a treated LRTI, having received antibiotics for more than 48 hours, or selected HIV-positive (<12 months old, uncontrolled or requiring admission) were excluded as written in the protocol. |
| Analysis Population | FAS | All baseline eligible participants. |
| Treatment conditions | Routine care ("Control") vs. use of LUS for treatment decision ("Intervention") |  |
| Endpoint | Clinical failure by day 8 | Note: this endpoint includes deaths due to non-trauma. |
| Summary measure | Risk differences. We will also report odds ratios and risk ratios. |  |
| Handling of intercurrent events (IE) [1] |  |  |
| Did not follow treatment assignment according to protocol | "Treatment policy" approach | The IE are defined as 1) not prescribing antibiotics in the "Control" arm and 2) not following the recommendations of the LUS in the "Intervention" arm. This IE is ignored and considered part of the intention to treat strategy |
| Death due to trauma | "While alive" approach | If the endpoint is measured prior to the IE, individual included and endpoint as recorded. If the endpoint is not measured prior to the IE, will be included as **not** having experienced clinical failure. |

[1] Intercurrent events are post-randomization events that affect either the interpretation or existence of the outcome data associated with the clinical question of interest (missing data or LTFU is not in itself an intercurrent event; see instead section 6.3 Missing data).

**Table 2d.**
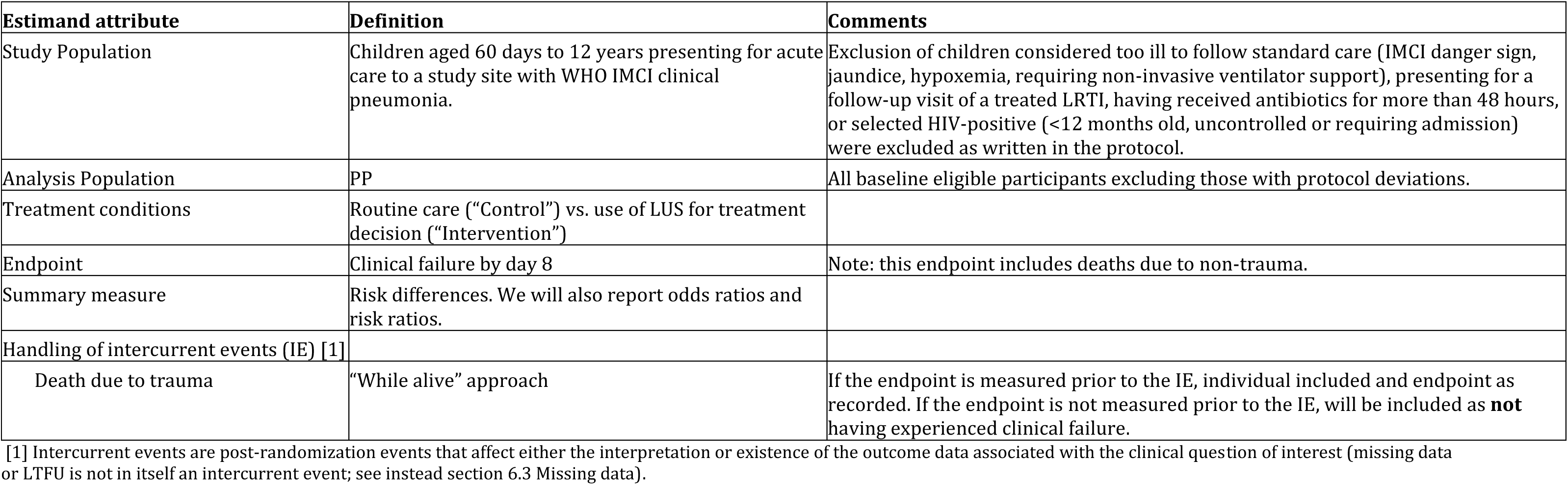
Estimand framework: second co-primary outcome of clinical failure using PP population [1].

| Estimand attribute | Definition | Comments |
| --- | --- | --- |
| Study Population | Children aged 60 days to 12 years presenting for acute care to a study site with WHO IMCI clinical pneumonia. | Exclusion of children considered too ill to follow standard care (IMCI danger sign, jaundice, hypoxemia, requiring non-invasive ventilator support), presenting for a follow-up visit of a treated LRTI, having received antibiotics for more than 48 hours, or selected HIV-positive (<12 months old, uncontrolled or requiring admission) were excluded as written in the protocol. |
| Analysis Population | PP | All baseline eligible participants excluding those with protocol deviations. |
| Treatment conditions | Routine care ("Control") vs. use of LUS for treatment decision ("Intervention") |  |
| Endpoint | Clinical failure by day 8 | Note: this endpoint includes deaths due to non-trauma. |
| Summary measure | Risk differences. We will also report odds ratios and risk ratios. |  |
| Handling of intercurrent events (IE) [1] |  |  |
| Death due to trauma | "While alive" approach | If the endpoint is measured prior to the IE, individual included and endpoint as recorded. If the endpoint is not measured prior to the IE, will be included as **not** having experienced clinical failure. |

[1] Intercurrent events are post-randomization events that affect either the interpretation or existence of the outcome data associated with the clinical question of interest (missing data or LTFU is not in itself an intercurrent event; see instead section 6.3 Missing data).

Secondary outcomes will be analyzed using only the FAS with a “treatment policy” approach. The estimand framework for the secondary binary outcomes is the same as for the primary outcomes (Table 2a-2d).

## 5. Study population

### 5.1 Screening

Every consecutive pediatric patient (60 days – 12 years) attending an acute outpatient department of a study site will be screened for participation in the study. Only patients meeting all the inclusion and none of the exclusion criteria will be recruited.

### 5.2 Eligibility

Screening/eligibility data will be summarized in a CONSORT flowchart, showing the total number of people screened and the reasons for screening failures as per the eligibility criteria listed below. See template in section 8.

**Inclusion Criteria:**

Children aged 60 days to 12 years presenting for acute care to a study site with WHO IMCI clinical pneumonia (and derived from IMCI for children above 5 years), defined as:

1. Cough **OR** Difficulty Breathing **AND**,
2. One of the below:

2a. Fast breathing (tachypnoea)

- ≥ 50/minute (2–12 months)
- ≥ 40/minute (1–5 years)
- ≥ 25/minute (5-12 years)
**OR**
2b. Lower chest wall indrawing

**Exclusion Criteria (these will apply for all children):**

1. Presenting for repeat visit/follow-up of a treated lower respiratory tract infection (index illness / non-acute) or enrolled in the study within the preceding 28 days.
2. Currently taking antibiotic treatment for more than 48 hours at the time of enrolment
3. WHO IMCI danger signs (*inability to drink/breastfeed, vomiting everything, convulsions with this illness, lethargy/unconscious*) : will also be applied for children ≥5 years of age
4. Presence of jaundice
5. Hypoxaemia with SpO2<88%
6. SpO2<90% (*or country-specific / altitude-adjusted thresholds*):

• With signs of severe respiratory distress (*such as nasal flaring, grunting, etc.*)
**OR**
• In children < 6 months
7. Requiring non-invasive ventilatory support (*i.e., high-flow, bi-PAP, CPAP*)
8. Underlying disease associated with increased risk of severe pneumonia or pneumonia of unusual aetiology *(e.g., WHO acute malnutrition requiring antibiotics as per local guidelines, severe immunodeficiency*)
9. HIV positive participant that is either:

• less than 12 months old,
**OR**
• requiring admission for this illness,
**OR**
• known to be uncontrolled on treatment (with a documented VL >1000c/ml in the previous 6 months)
10. Caregiver unavailable at the time of enrolment, or unwilling, to provide informed consent.

### 5.3 Recruitment

We will present screening and randomization graphically over time. The CONSORT flowchart will include the numbers of participants randomized by arm. Enrolment will be presented by the age group, site, and country.

The planned breakdown of participant enrolment per country is as follows:

- Tanzania – 3 sites and 2000 participants
- Senegal – 1 site and 500 participants,
- South Africa – 3 sites and 1000 participants. Recruitment is expected to take a maximum of 24 months.

### 5.4 Withdrawal/follow-up

The CONSORT flowchart will summarize each of the scheduled assessment time points (Day 1, Day 8 and Day 29) by study arm. The number of children withdrawn in between follow-ups will also be summarized and reasons provided if available.

At enrollment, the participant’s primary caregiver will be asked to provide the contact of a next-of-kin to assist in collection of follow-up information in the unfortunate event of unavailability, death, or incapacitation of the primary caregiver.

On D8, a telephone follow-up will be conducted with the caregiver to assess for current health status, evidence of clinical failure, clinical course since enrolment, and antibiotic use. Figure 1 provides a detailed algorithm of procedures to be followed. If caregivers cannot be reached by phone, other strategies detailed in the Manual of Operations will be implemented to reach these caregivers, including home visits, and establishing contact through community stakeholders (community leaders, community health workers).

**Figure 1:**
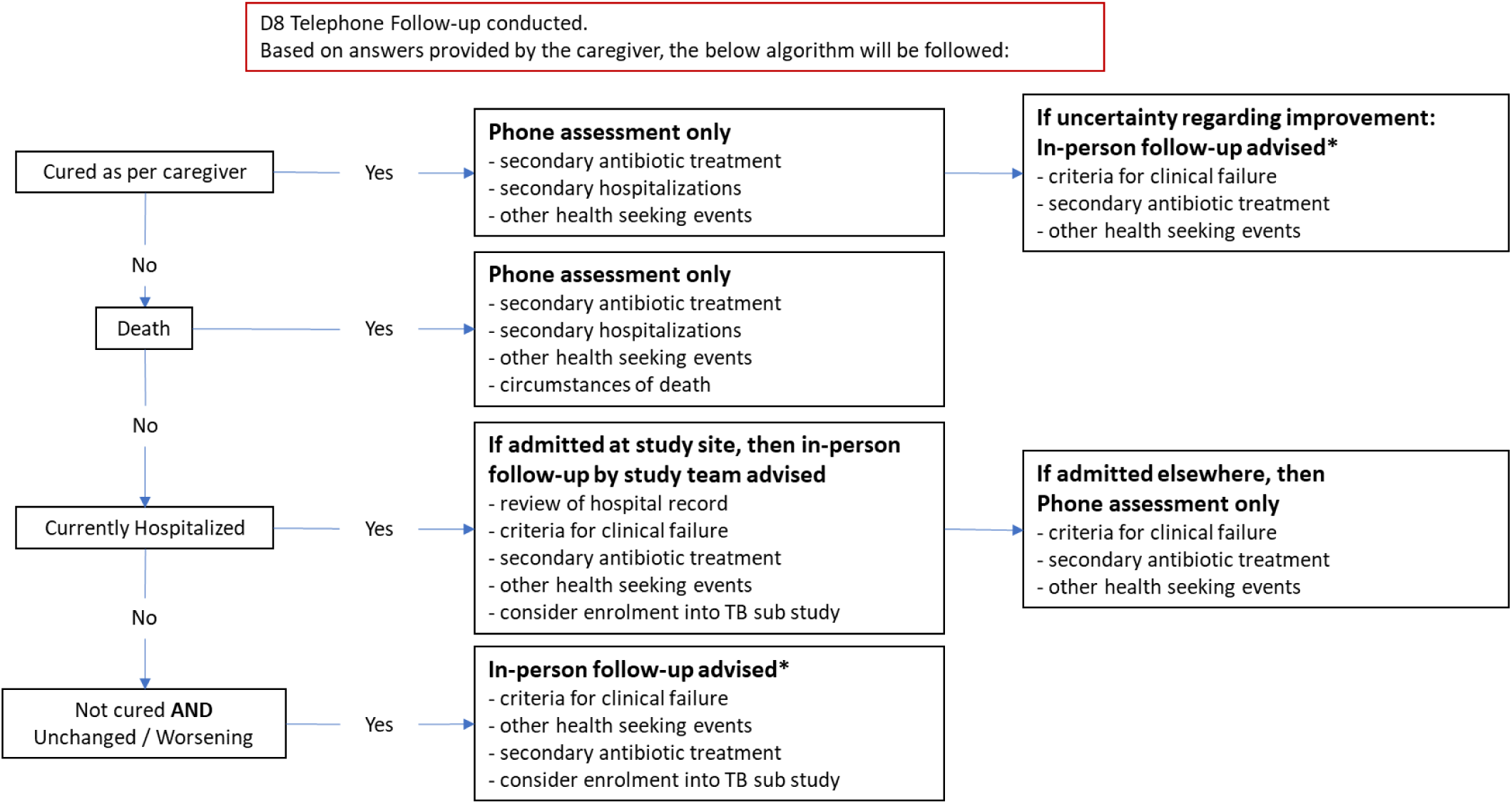
Follow-up procedures for those reached during follow-up.

On D29, a telephone follow-up will be conducted to assess for repeat health seeking events for a respiratory illness, hospitalization, history of starting TB treatment, and death.

Participants who are not reachable within D29 window will be declared as lost to follow-up.

### 5.5 Baseline patient characteristics

Baseline characteristics will be summarized by randomized group and age group, using medians and interquartile ranges for continuous variables and numbers and percentages for categorical variables. Summary statistics will be of non-missing values, with the number (%) of missing values given if data are not complete. The table will be solely descriptive and no formal testing of baseline characteristics across randomized groups [8–10]. A shell table showing the variables and categories is included in section 8.

## 6. Analysis

As per the protocol, all analyses will be done by the trial statistician. A senior statistician will review the statistical reports and validate the analysis, following a risk-based approach.

Analyses will follow CONSORT guidelines, including extensions for non-inferiority trials [10–12]. Analyses will include all follow up to the date of data lock. Analyses will be performed and reported overall and by age group.

Percentages will be reported to zero decimal places, unless <0.5% when they will be given to one decimal place. Means, standard deviations, medians, and interquartile ranges will be reported to one decimal place.

### 6.1 Outcome definitions

Here we define outcomes for analysis. The definitions are based on protocol defined assessment window, as shown in Table 1. For example, if an endpoint is defined by Day 8, any assessments that contributes to that endpoint done within 12 days from randomization will be included in the definition (8 days + 4 window days). The date of randomization (randdate) is the automated date generated when the randomization form is opened in the randomization database.

The denominators for the outcomes are based on the analysis populations as indicated in section 4.3, unless otherwise specified.

Data validation checks (detailed in the Data Review Log) will be performed throughout the study to ensure that outcomes are defined using reliable and consistent data. If data changes are required as a result of the checks, these will be done before database lock. In the event of unresolved conflicting information recorded on different forms, any evidence of the occurrence of the event will be counted for the outcomes definition.

#### 6.1.1 Primary outcomes

**Co-Primary Endpoints**

i) Antibiotic prescription on D1 (yes/no). This information comes **solely** from the Health Seeking, Medication and Antibiotics form. Several clinicians may see the patient on day 1 (e.g. a physician at the health facility and an admitting team). The Health Seeking, Medication and Antibiotic form will be filled in based on the management decision of the first clinician evaluating the patient.

- If for the Day 1 – Clinical History (Meds. before enrollment) (cm_visit=1) or Day 1 – Routine Clinician’s Prescription (cm_visit=2), the answer to the following question ‘Type of entry’ is ‘Antibiotic’ (cm_type_1=1), then the child will meet the definition of this endpoint.
- When route is topical, rectal, or inhalation (cm_route=4,5,6), this will not be considered as an antibiotic prescription.
ii) Clinical failure any time after randomization or before D8 (yes/no). The child will meet the definition of this endpoint if any of the following conditions are true:

- WHO IMCI danger signs captured on Physical Exam form after randomization and before day 8 window (event_name ≠ day_1_arm_1 AND pe_date-randdate≤12 days): (inability to drink/breastfeed (sym_imci_danger_signs___1=1), vomiting everything (sym_imci_danger_signs___2=1), convulsions with this illness (sym_imci_danger_signs___3=1), lethargy/unconscious (sym_imci_danger_signs___4=1)
- New or worsening severe respiratory distress captured on Physical Exam form after randomization and before day 8 window (event_name ≠ day_1_arm_1 AND pe_date-randdate≤ 12 days) (any one of the following):

◦ Nasal flaring (pe_sym_respidistress___2), grunting (pe_sym_respidistress___3), head nodding (pe_sym_respidistress___4), tracheal tug (pe_sym_respidistress___5), intercostal recessions (pe_sym_respidistress___ 6), observed apnoea (pe_sym_respidistress___7) AND pe_respiratory_distress=1 or 2)
◦ Severe chest indrawing (pe_chest_indrawing=2))
◦ Stridor at rest (pe_stridor=1)
- Secondary Hospitalization (defined as a hospitalization occurring after discharge from in-patient admission or outpatient visit) related to a deterioration of the presenting complaint on D1 will come from any one of the following sources:

◦ Post Clinical Review: Referred from ED/OPD for admission today (for this illness) (exit_disposition=2) AND Admitted In-ward Follow-up form: admitted after D1 (inward_hosptype=3) AND by D8 window (inward_admissiondate-randdate≤12 days).
◦ Phone & General Follow-up form: a suitable person was reached (fusp_phone_result=1) AND admitted after D1 (fusp_hospitalization=3) AND related to the initial illness (fusp_ illness_hospitalization=1 or 98) AND admission date by D8 window (fusp_admission_date-randdate≤12 days).
- Change in level of care (including on day 1) (e.g., admission to intensive care unit, transfer to higher level of care) will come from any one of the following sources:

◦ Post Clinician Review: exit_disposition = 5 AND exit_interview_date-randdate≤12 days.
◦ Admitted In-ward Follow-up form: inward_level_care_change=1,2, or 3 AND inward_follow_up_date-randdate≤12 days.
- Need for respiratory support (e.g., high flow nasal cannula, CPAP): vsm_spo2_mech=2, 3, 4, 5 or 6 AND vsm_date-randdate>1 day AND vsm_date-randdate≤12 days; captured on Vitals form.
- Death due to any medical cause (i.e., except trauma) will come from:

◦ End of Study form: eos_reason=2 AND eos_death_medicaltrauma≠2 AND eos_death_date-randdate≤12 days
- Report from the caregiver of non-resolution/worsening of illness at D8 will come from any one of the following sources:

◦ Phone Follow-up form: fusp_phone_result=1 AND fusp_phone_ch_outcome =3 or 4 AND fusp_actual_date-randdate≤12 days.
◦ In-person follow-up: inperson_visit_reason=1 AND inperson_patient_outcome=3 or 4 AND inperson_fu_actual_date-randdate≤12 days

#### 6.1.2 Secondary outcomes

iii) The proportion of children prescribed an antibiotic by D8 window (yes/no) will come from any one of the following sources:

- Anyone meeting the criteria for antibiotics on D1 (primary endpoint) also meets the criteria for this endpoint (see above 6.1.1 i).
- Health Seeking, Medication and Antibiotics form: if they were prescribed antibiotic (cm_type_1=1) and the start date of the prescription is before D8 window (cm_cmstdate-randdate≤12 days), then the child will meet the definition of this endpoint.

◦ When route is topical, rectal, or inhalation (cm_route=4,5,6), this will not be considered as an antibiotic prescription.
- Caregiver says they took antibiotics on the day 8 phone follow-up:

◦ Phone & General follow-up form: fusp_phone_result=1 AND (fusp_phone_atb_taken= 1 OR fusp_atbno>0) AND fusp_actual_date-randdate≤12 days. This should provoke the Health Seeking form to be completed but will be counted as reaching the endpoint without this confirmation.
iv) The proportion of adverse drug reactions related to routine antibiotic treatment by D8 window (i.e., Anaphylactic reaction, severe diarrhoea, or generalised severe rash) will come from any one of the following sources:

- Medical & adverse event form: ae_name_class==1 (ADR) AND ae_date_onset-randdate≤12 days
- Physical exam form: pe_drugreaction 1=1, 2=1 or 3=1 AND pe_date-randdate≤12 days
- Phone & General follow-up form: fusp_phone_result=1 AND fusp_inward_drugreaction___1=1, 2=1, or 3=1 AND fusp_actual_date-randdate≤12 days
- Health seeking, medication & antibiotics form : cm_visit = 9 (ADR) AND cm_type_1=1 AND cm_cmstdate-randdate≤12 days
v) The proportion of patients cured (defined as caregiver reported recovery from illness) will come from any one of the following sources:

- By D8 (Phone Follow-up form: (fusp_phone_result=1 AND fusp_phone_ch_outcome =1 AND fusp_actual_date-randdate≤12 days); denominator is all children as indicated in Table 2b (FAS).
- D29 (Phone Follow-up form: (fusp_phone_result=1 AND fusp_phone_ch_outcome =1 AND fusp_actual_date-randdate≤33 days); denominator is children as indicated in Table 2b (FAS) AND not cured at D8.
vi) The proportion of patients admitted to hospital on D1 for any reason will come from any one of the following sources. In the event of conflicting information, any evidence of the occurrence of the event will be counted.

- Phone & General Follow-up form: fusp_phone_result=1 AND fusp_hospitalization=2
- Admitted In-ward follow-up form: inward_hosptype=2
vii) The proportion of patients admitted to hospital for any reason by D8 window will come from any one of the following sources:

- Anyone admitted on Day 1 as specified above.
- Anyone admitted after Day 1 and before Day 8:

◦ Admitted In-ward Follow-up form: inward_hosptype=3 AND inward_admissiondate-randdate≤12 days.
◦ Phone & General Follow-up form: fusp_phone_result=1 AND fusp_hospitalization=3 AND fusp_admission_date-randdate≤12 days.
viii) The duration of the D1 inpatient admissions (censored at D29) will come from any one of the following sources:

- Phone & General Follow-up form: for those with fusp_hospitalization=2, the duration=min(fusp_discharged_date – randdate, 33)
- Admitted Inward Follow-up form: for those with inward_hosptype=2, the duration=min(inward_discharge_date – randdate, 33)

The denominator includes all children from FAS admitted on D1.

ix) The duration of any inpatient admissions starting between D1 (included) and D8 (censored by D29) (i.e., secondary hospitalizations but does not need to be related to the initial illness) will come from any one of the following sources:

- Anyone admitted on Day1 as specified above.
- Anyone admitted after Day1 and before Day8:

◦ Phone & General Follow-up form: for those with fusp_hospitalization=3 and fusp_admission_date-randdate≤12days, the duration=min(fusp_discharged_date – fusp_admission_date, 33)
◦ Admitted Inward Follow-up form: for those with inward_hosptype=3 and inward_admissiondate≤12 days, the duration=min(inward_discharge_date – inward_admissiondate, 33)

The denominator includes all children from FAS admitted between D1 (included) and D8.

x) The proportion of patients undergoing a non-study related diagnostic test (including CXR, blood tests, urine tests, microbiological assays) on D1 and through D8 for those admitted on D1; captured on the Routine Investigations form.

- on D1:

◦ Routine tests done (rout_done=1) AND on day 1 (rout_date=randdate) AND (POC tests (hemoglobin (rout_hb_test_yn=1) OR glucose (rout_gluc_test_yn=1) OR malaria (rout_malaria_test=0, 1, 3) OR HIV (hist_hiv_rapid_test=0, 1, 3) OR urine LAM (rout_lam_test=0, 1)) OR routine tests (rout_samples___1=1, 2=1, 3=1, 4=1) OR chest x-ray (rout_cxr=1))
- Admitted on D1, tests conducted up until D8; denominator is all children in the FAS who were admitted on D1:

◦ Routine tests done (rout_done=1) AND before Day 8 (rout_date-randdate≤12) AND (hemoglobin (rout_hb_test_yn=1) OR glucose (rout_gluc_test_yn=1) OR POC tests (rout_samples___1=1, 2=1, 3=1, 4 =1) OR chest x-ray (rout_cxr=1))
xi) The proportion of deaths due to any cause by D29:

- End of Study form (eos_reason==2 AND eos_death_date-randdate≤33 days);
xii) The proportion of participants with unscheduled health seeking events for any cause and/or hospital admission for any cause since D8 follow-up (at D29 follow-up):

- Phone & General Follow-up form: Reached caregiver (fusp_phone_result=1) AND between day 8 and 29 (fusp_actual_date-randdate>12 days AND fusp_actual_date-randdate≤33 days) AND (additional visits to the facility (fusp_facvisitno>0) OR outpatient visits (fusp_outpvisitno>0) OR pharmacy visits (fusp_pharmvisitno>0) OR visits to traditional healer (fusp_tradvisitno>0) OR hospitalization for any reason (fusp_hospitalization=3 AND fusp_admission_date-randdate>12 days)).

Secondary objectives for the TB sub-study, cost-effectiveness, as well as exploratory endpoints will be further described in a separate document by the respective study teams.

### 6.2 Analysis methods

Outcomes will be described by arm using summary statistics. Outcomes will also be summarized within randomized arm by age group and country.

In particular, counts and percentages will be used for binary variables and reported with one decimal place; mean and standard deviation (SD) or median (IQR) for continuous variables depending on their distribution. Mean and median will be displayed with one decimal place and SD and IQR will be presented with two decimal places.

In case the same child is enrolled multiple times during the study enrollment period, only the first enrollment will be included in the primary analysis of primary and secondary outcomes.

#### 6.2.1 Primary outcome analysis

The primary analysis for this study will be the comparison of antibiotic prescription and clinical failure rates between the two study arms. For the primary outcome, we will summarize the numbers and percentages of participants included in the analysis, and those experiencing intercurrent events, as per Tables 2a-2d. All primary analysis results stem from multiple imputation as specified in Section 6.3.

The analysis of the first primary outcome, the proportion of children receiving antibiotics on D1, will be assessed for superiority using both the FAS and the PPS (Table 2a-Table2b). A modified Poisson regression with an identity link and robust standard errors will estimate the risk difference and respective 95% confidence interval [13]. In the case of convergence issues, we will use marginal standardization to obtain the risk differences [14]. In addition to the absolute effect measure of risk differences, we will report the relative effect of the intervention with odds ratios and risk ratios as recommended by the CONSORT statement [15]. Odds ratios will be estimated using logistic regression with logit link and risk ratios using a modified Poisson regression with a log link and robust standard errors. All effect estimates will be presented with 95% confidence intervals. All models will be adjusted for age group and site, the pre-specified randomization stratification factors [16].

The second primary outcome, the proportion of children with clinical failure by D8, will first be evaluated for non-inferiority using a one-sided CI approach. A figure illustrating the 95% CIs of the risk difference and the non-inferiority margin will be presented.

Primary analyses for the non-inferiority comparison will be performed on both the PPS and FAS (Table 2c-Table2d). If the upper bound of a two-sided 95% confidence interval for the risk difference does not include the non-inferiority margin of 2.5%, then the intervention will be considered non-inferior. If clinical failure in the intervention group is found to be non-inferior to the control, then we will assess for superiority using the FAS [6]. If the upper limit of the two-sided 95% confidence interval is less than 0, the intervention will be considered superior with respect to clinical cure. As this corresponds to a closed test procedure, no adjustment for multiple testing is needed [6]. The estimates and CIs will be obtained from a modified Poisson regression model and adjusted for the stratification factors of site and age group (as for the first primary outcome).

For the intervention to be considered successful, all null hypotheses must be rejected. In other words, the intervention should be shown to significantly reduce antibiotic prescription and be non-inferior with respect to clinical failure as compared to the control. Secondary outcomes should only be interpreted under these conditions.

##### 6.2.1.1 Planned sensitivity analysis

The following sensitivity analysis are planned:

- For both co-primary outcomes, we will additionally adjust models for pre-specified prognostic baseline characteristics as this has been shown to improve efficiency [9, 16–17].

Models for antibiotic prescription will be adjust for:

◦ Age groups (below 6 months, 6 to 12 months, 12 to 24 months, 2 to 5 years, above 5 years) [scr_child_dob, calc_estmated_dob]
◦ Month of enrollment (Jan-Mar, Apr-Jun, Jul-Sep, Oct-Dec) [randdate]
◦ Past medical history of asthma (yes, no) [hist_chronic_dx_type=1]
◦ Past medical history of heart problem (yes, no) [hist_chronic_dx_type=2]
◦ Past medical history of cerebral palsy (CP) or neurological problem (yes, no) [hist_chronic_dx_type=3]
◦ Past medical history of prematurity (< 37 weeks) (yes, no) [hist_chronic_dx_type=4]
◦ Malaria mRDT result (positive, negative, unknown) [rout_malaria_test]
◦ SpO2 at baseline (88% to 92%, >92%) [vsm_spo2]
◦ Ear ache/discharge at baseline (yes, no) [enrol_symp_ear]
◦ URTI at baseline (yes, no) [enrol_symp_urt]
Models for clinical failure will be adjusted for:

◦ Age groups (below 6 months, 6 to 12 months, 12 to 24 months, 2 to 5 years, above 5 years) [scr_child_dob, calc_estmated_dob]
◦ Month of enrollment (Jan-Mar, Apr-Jun, Jul-Sep, Oct-Dec) [randdate]
◦ Past medical history of asthma (yes, no) [hist_chronic_dx_type=1]
◦ Past medical history of heart problem (yes, no) [hist_chronic_dx_type=2]
◦ Past medical history of cerebral palsy (CP) or neurological problem (yes, no) [hist_chronic_dx_type=3]
◦ Past medical history of prematurity (< 37 weeks) (yes, no) [hist_chronic_dx_type=4]
◦ Past medical history of kidney problem (yes, no) [hist_chronic_dx_type=5]
◦ Malaria mRDT result (positive, negative, unknown) [rout_malaria_test]
◦ SpO2 at baseline (88% to 92%, >92%) [vsm_spo2]
◦ Weight for high score (above 3+, +3 to +2, +2 to 0, 0 to −2) [vsm_anthro_wfh]
◦ MUAC (<11.5, 11.5 to 12.5, >12.5) [vsm_muac]
◦ CRP at baseline (<10, 10 to 40, 40 to 80, >80) [biob_edta250_crp_result]
- Very early clinical failure cases will most likely be due to miss-classification of the severity of illness or rapid deterioration, which we hypothesize would be observed independent of arm assignment. However, as the number and potential impact of such cases is not known, a sensitivity analysis will be conducted for both co-primary outcomes excluding from the FAS those participants who experience clinical failure on Day1.
- A sensitivity analysis on the PPS, excluding those who did not adhere to the protocol (see 4.2) will be conducted for both co-primary outcomes.
- A complete cases sensitivity analysis, excluding those with missing co-primary outcomes, will be performed.
- A sensitivity analysis on all enrollment episodes, including multiple enrollments for the same child, will be conducted on both co-primary outcomes. Generalized estimating equations (GEE) for modified Poisson and logistic regression will be used to estimate absolute effect measure (risk difference) and relative effect measures (risk ratio and odds ratio) respectively. Children will be modeled as clusters to account for correlated observations.

##### 6.2.1.2 Planned subgroup analysis

The study was powered for children under 5 year of age (see 3.4) as this is the subgroup of greatest interest based on current IMCI framework. Effect modification of age will be assessed by incorporating an interaction between arms and age (under 5 and 5-12 years). Stratified model results will be presented for children under 5 and children 5-12 regardless of the significance of the interaction term. Effect estimates of the co-primary outcomes will be presented as for the primary outcome analysis.

#### 6.2.2 Secondary outcomes analysis

The secondary outcomes are not part of the confirmative strategy but are intended to provide supportive evidence without confirmative conclusions in the case where the co-primary outcomes are all positive.

All secondary binary outcomes (except those explicitly mentioned below) will be tested for superiority using unadjusted models in the FAS population (Table 2a and 2c; see details in 6.1.2) as with the primary outcome(s) and summarized with risk differences, risk ratios, odds ratios and 95% Wald confidence intervals.

Rare outcomes such as death will be compared across arms using non-parametric or small sample tests.

Hospitalization is only expected in a subset of participants and the length of stay is not expected to be normally distributed. The secondary outcomes of duration of hospitalization will be tested non-parametrically using a Wilcoxon rank-sum test.

#### 6.2.3 Safety outcomes

Safety data will be collected, reported, and recorded as described in the protocol. Adverse events (AEs) and serious adverse events (SAEs) will be summarized by trial arm.

### 6.3 Missing data

Missing baseline and outcome data will be summarized by study group.

As the co-primary outcome of antibiotic prescription is recorded on Day 1 (same day as randomization), we do not expect this information to be missing. If however more than 3% of individuals are missing this outcome data, we will impute the outcome using a multiple imputation approach under the assumption of data missing at random. We will used chained equations with 50 imputed datasets [19,20]. In the imputations, we will use baseline variables plus follow-up data (Table 3). We will perform the imputation separately by randomized arm [21]. Results are combined by Rubin’s rule.

**Table 3.**
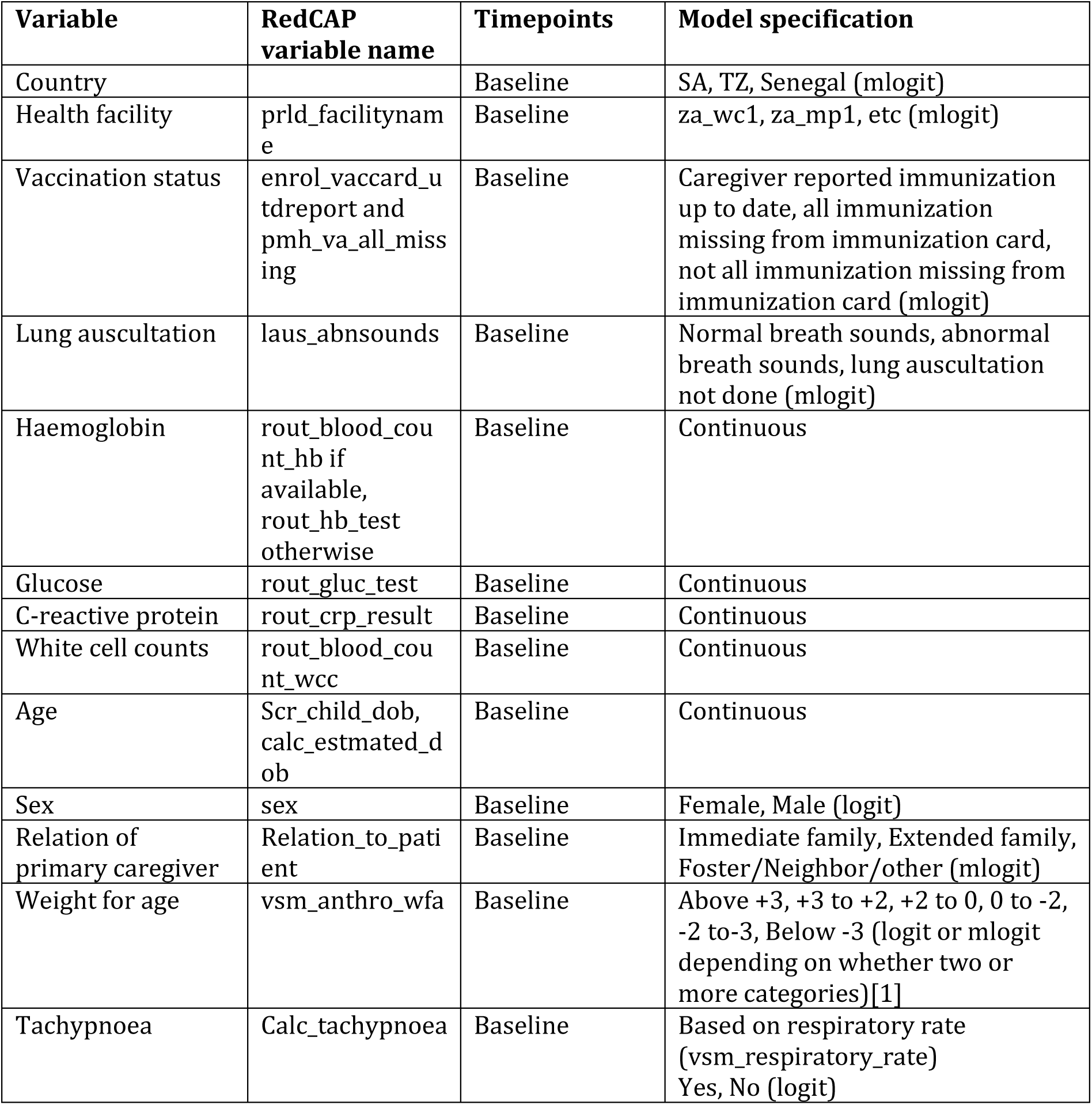
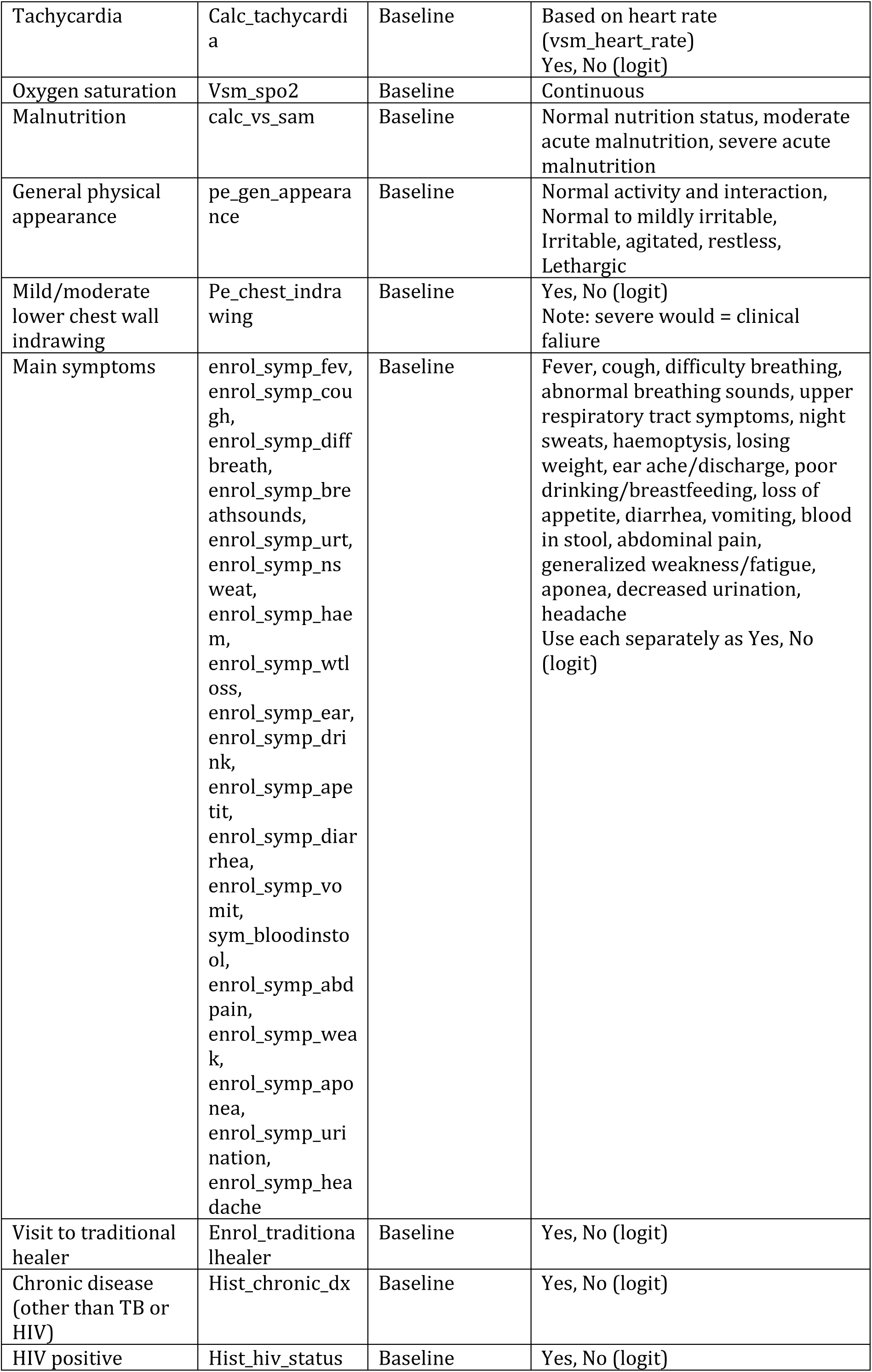
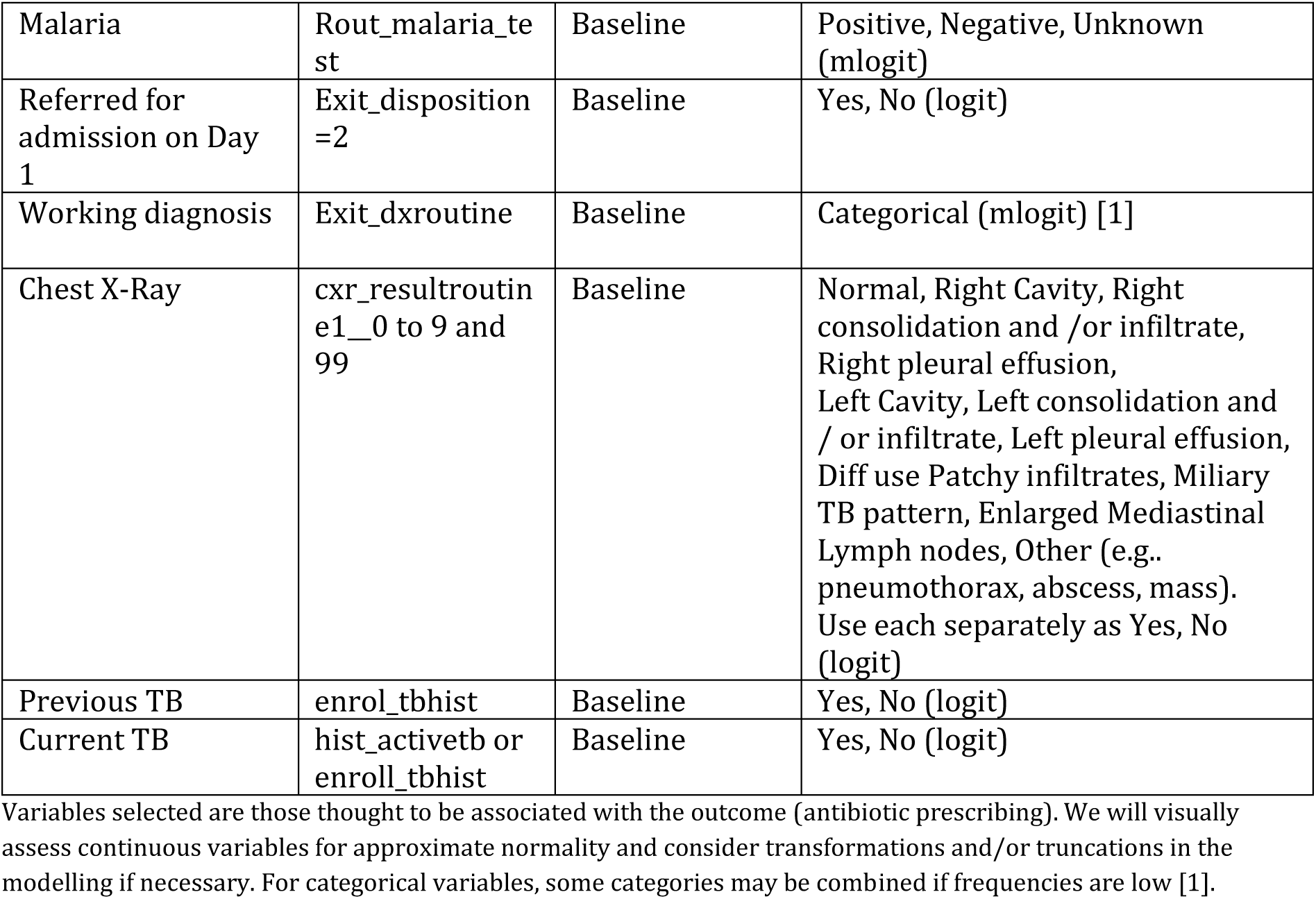
Variables to be included in multiple imputation of co-primary endpoint of antibiotic prescribing.

Note that the secondary outcome of antibiotic prescription by day 8 will be imputed as per the co-primary outcome described above.

For the co-primary outcome of clinical failure, we will use multiple imputation to account for data missing due to not being reached for the Day 8 follow-up or withdrawal of consent, under the assumption of data missing at random. We will used chained equations with 50 imputed datasets [19,20]. In the imputations, we will use baseline variables plus follow-up data (Table 4). We will impute missing values of the individual elements of clinical failure (see 6.1.1), and then use these values to determine whether the outcome of clinical failure by Day 8 was met. We will perform the imputation separately by randomized arm [21]. Results are combined by Rubin’s rule.

**Table 4.**
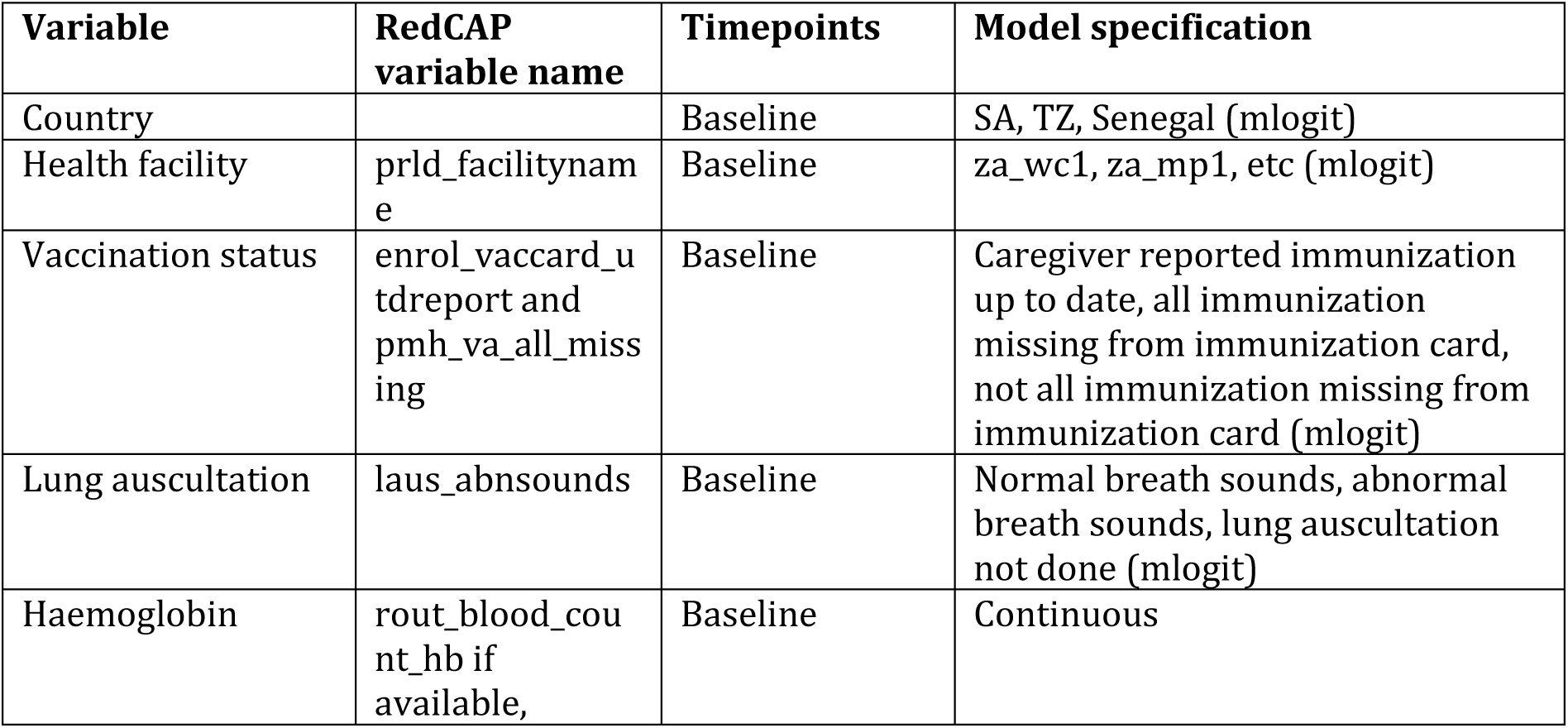
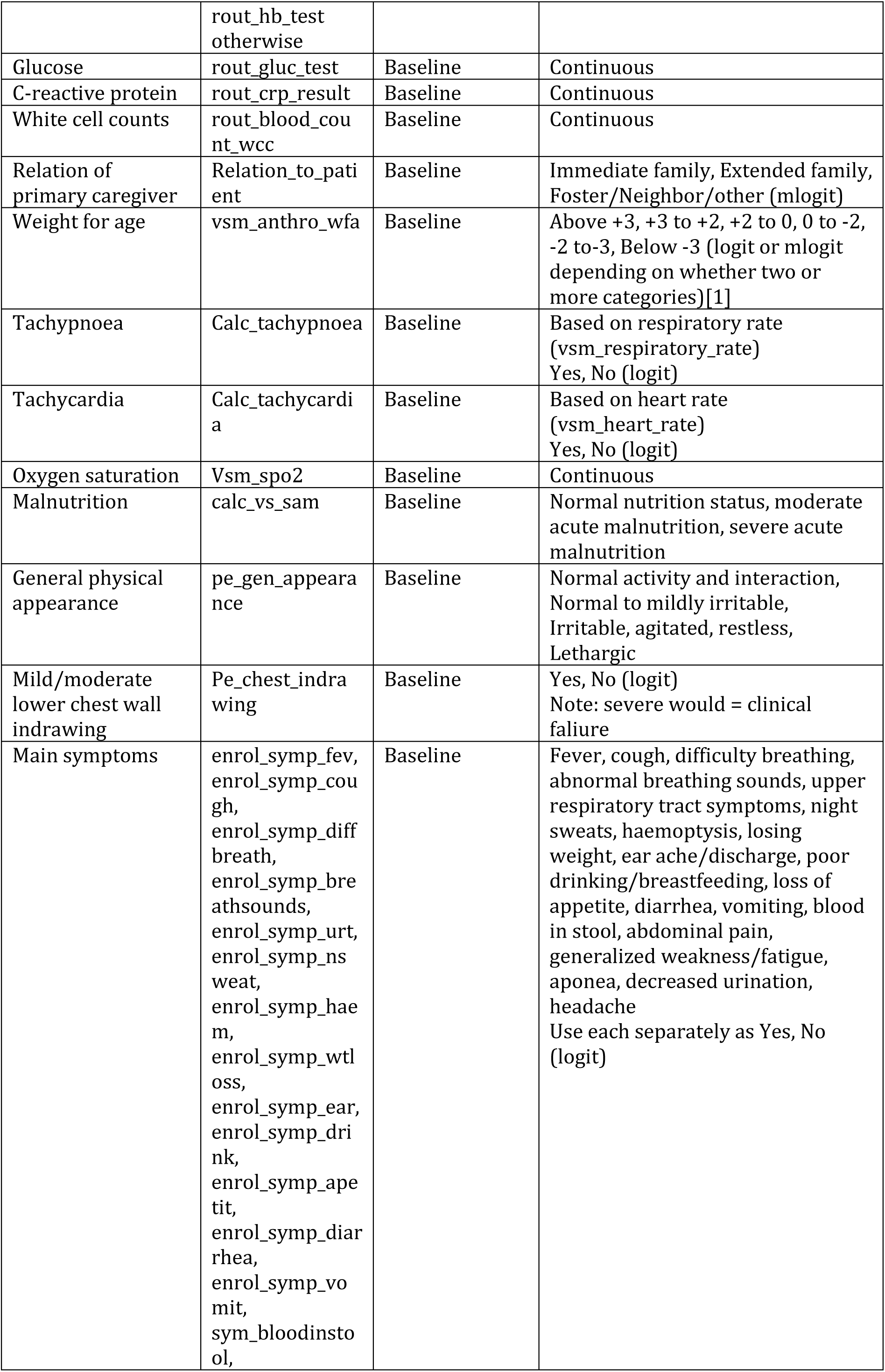
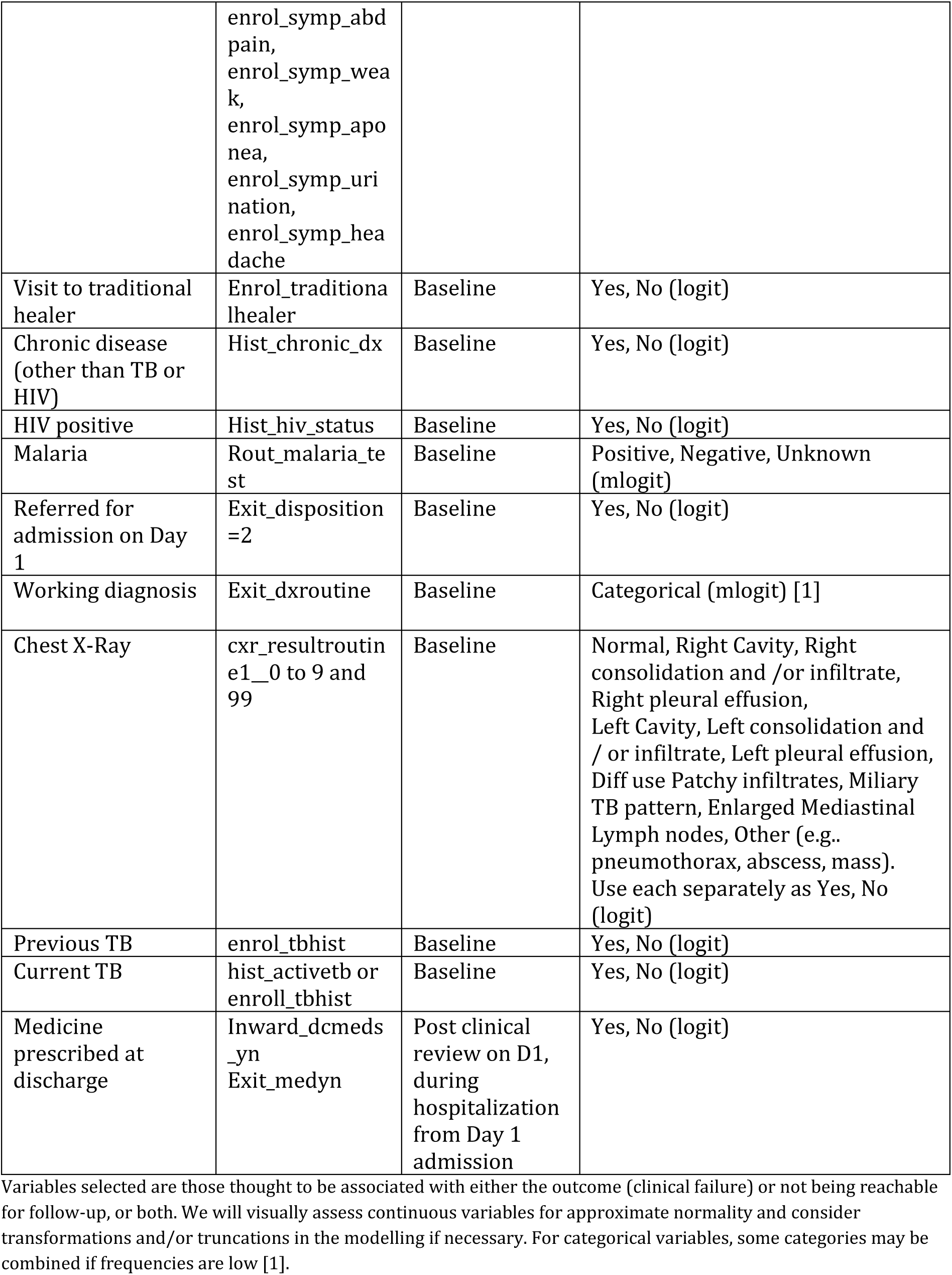
Variables to be included in multiple imputation of the individual elements of co-primary endpoint of clinical failure.

For comparison purposes, in addition we will perform complete case analyses for the co-primary outcomes, including only the participants with outcome data. This would be a sensitivity analysis under the assumption of data missing completely at random.

Other secondary outcomes (not specifically mentioned above) to be imputed (as described above and in Table 4) are duration of Day 1 admissions, cure by day 29, and death by day 29.

All other secondary outcomes will be analyzed with a complete case analysis.

### 6.4 Additional analyses

Analyses for sub-studies and cost-effectiveness will be described elsewhere.

### 6.5 Statistical software

All analyses will be done using R (version 4.1.2 or higher, the R Foundation for Statistical Computing) or Stata (version 18 or higher, Stata Corporation, Austin/Texas, USA) and use version control software (Git). The software and version used for each analysis will be documented in the statistical report.

## Data Availability

not applicable

## Abbreviations

AE: Adverse Event
CI: Confidence Intervals
CPAP: Continuous Positive Airway Pressure
CRF: Case Report Form
CXR: Chest X-Ray
DMP: Data Management Plan
eCRF: electronic Case Report Form
EDC: Electronic Data Capture
ER: Emergency Room
FAS: Full Analysis Set
HIV: Human Immunodeficiency Virus
IDMC: Independent Data Monitoring Committee
IMCI: Integrated Management of Childhood Illness
IMCI-PLUS: IMCI-Paediatric Lung Ultrasound
ITT: Intention to Treat
LUS: Lung Ultrasound
LAusc: Lung Auscultation
LRTI: Lower respiratory tract infection
LTFU: Lost to Follow-Up
PPS: Per-protocol set
RCT: Randomised Controlled Trial
RSV: Respiratory Syncytial Virus
SAE: Serious Adverse Event
SSA: Sub-Saharan Africa
TB: Tuberculosis
WHO: World Health Organisation

## 9. Appendix A: sample size calculation

Assumptions related to the co-primary outcomes:

- Antibiotic prescription

◦ According to the IMCI framework, all children with IMCI pneumonia should receive antibiotics. However, due to non-compliance with recommendations or extenuating circumstances, we expect approximately 80% current prescription rate in the control arm.
◦ A reduction of ≥15% is considered clinically meaningful.
- Clinical/treatment failure

◦ A rough literature search of all RCTs relating to Paediatric Community acquired Pneumonia (CAP) in the last 10 years yielded approximately 280 articles, of which 35 appeared relevant. Below a summary of key takeaways:

▪ Most define a composite endpoint (“treatment/clinical failure”) whose definition differs between studies.
▪ Most studies that provide a rationale for the delta they used, explain a 1.5x margin as an acceptable margin to clinicians (i.e. 50% change) [25, 26].
▪ Some studies uses an interim analysis to adjust the sample size and the margin [25–28].
◦ Taking a more conservative approach, we explored the sample size calculation under the following assumptions:

▪ A proportion of clinical failures lower than 10%, such as 4-7%.
▪ Since our clinical failure definition combines clinical outcomes (like death/WHO danger signs etc.) and treatment failure (which is a less severe outcome than death or disease complications), a higher rate of failure may be acceptable/considered clinically non-inferior to a panel of Clinicians - e.g. 60-70%. So we have used the 1.5x as per the above-mentioned studies but also explored the other options of 60% and 70% change (labelled as 1.6x and 1.7x).
▪ The loss to follow-up of 5%.

**Table 5:**
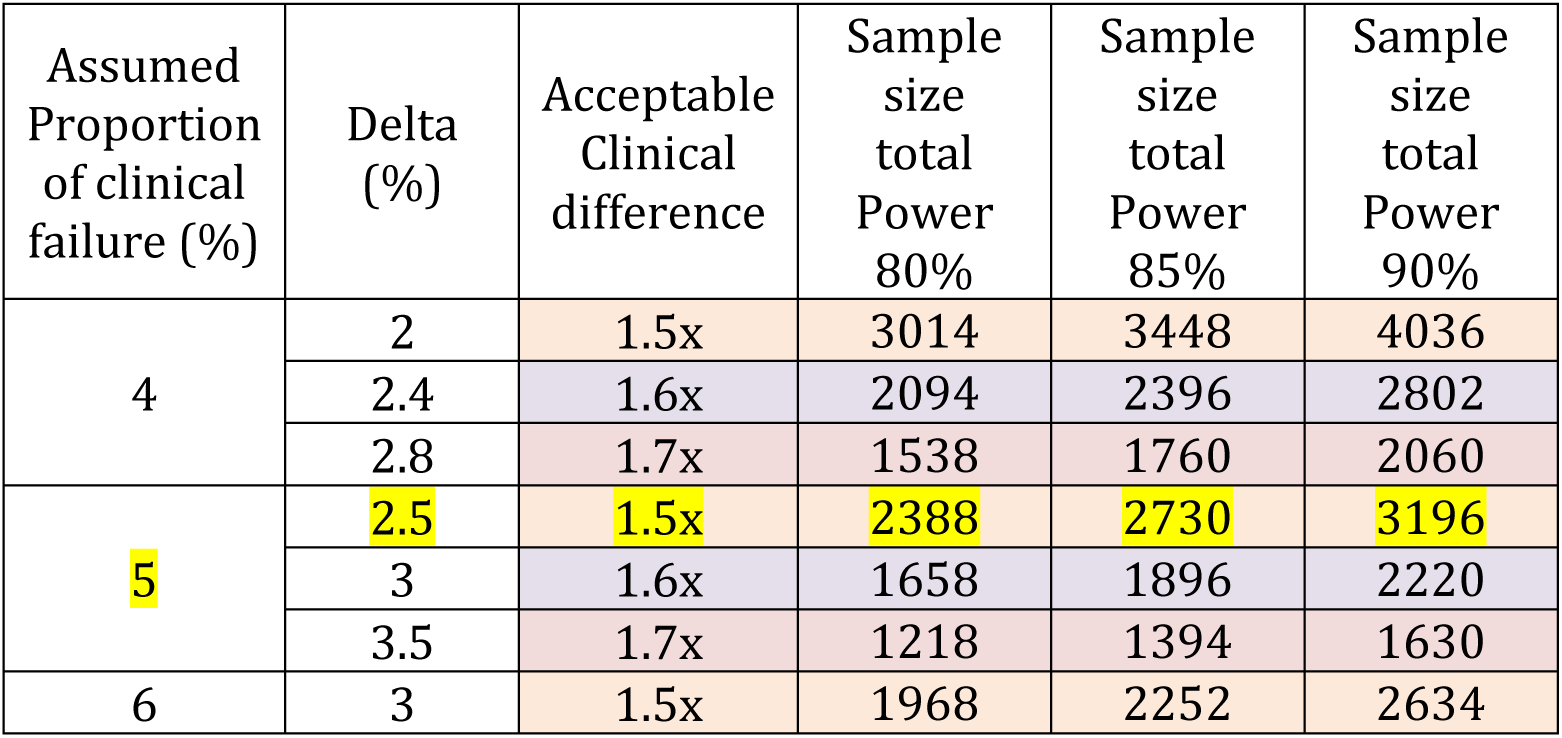
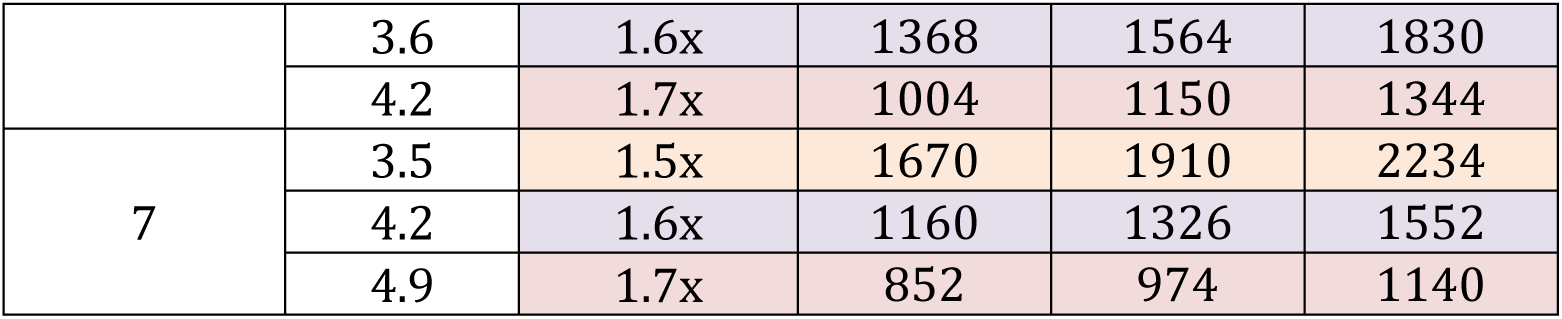
Sample size calculation for clinical failure rate under different assumptions.

After evaluating the options above, it was decided to assume the proportion of clinical failure in the control group as 5%, an acceptable clinical difference of 1.5x as a non-inferiority margin (therefore a delta of 2.5%), and a power of 85% (highlighted row). This led to the estimated sample size required across all sites of 2730 children (1365 per arm). This would equate to 1365 participants per group for the co-primary endpoints.

## 10. Appendix B: simulation studies

We performed 10000 Monte Carlo simulation runs, except only 1000 for power for futility (10.4), to evaluate different interim analysis scenarios. All simulations were performed using R version 4.4.1. Results of the simulations guided us to develop the strategy for the sample size re-calculation and conditional power assessment.

The scenarios tested are for an interim time point of 50% of the total planned patients (1365 per arm) with the following assumptions on the true antibiotic rate and clinical failure rate.

| Scenario | Outcome | Group | Value |
| --- | --- | --- | --- |
| "0.8/0.77" | Antibiotic rate | Control arm | 80% |
|  | Antibiotic rate | Experimental arm | 77% |
|  | Clinical failure rate | Control arm | 1%, 2.5%, 5%, 7%, 10% |
|  | Clinical failure rate | Experimental arm | 1%, 2.5%, 5%, 7%, 10% |
| "0.8/0.7" | Antibiotic rate | Control arm | 80% |
|  | Antibiotic rate | Experimental arm | 70% |
|  | Clinical failure rate | Control arm | 1%, 2.5%, 5%, 7%, 10% |
|  | Clinical failure rate | Experimental arm | 1%, 2.5%, 5%, 7%, 10% |
| "0.8/0.65" | Antibiotic rate | Control arm | 80% |
|  | Antibiotic rate | Experimental arm | 65% |
|  | Clinical failure rate | Control arm | 1%, 2.5%, 5%, 7%, 10% |
|  | Clinical failure rate | Experimental arm | 1%, 2.5%, 5%, 7%, 10% |

We simulated independent binary co-primary endpoints. For the first co-primary endpoint (superiority setting) we calculated the upper 95% Wald confidence interval of a risk difference and concluded superiority if the 95% confidence interval excluded zero. For the second co-primary endpoint (non-inferiority setting) we calculated the upper 95% Wald confidence interval of a risk difference and concluded non-inferiority if the upper 95% confidence limit was below the non-inferiority margin of *δ* = 1.5*pa* − *pa*, where *pa* is the pooled clinical failure rate at the interim analysis.

In addition, as the absolute non-inferiority margin would change at the interim analysis, we assessed the type-I error [24] assuming an antibiotic rate of 80% in the control arm and experimental arm.

### 10.1 Sample size re-estimation

We calculated the re-estimated power for the co-primary under the sample size of the planned 1365 patients per arm. The dashed red line indicates the agreed power (85%) for the initial sample size calculation. Reading example (most right panel): In case of a true antibiotic rate of 80% in the experimental arm and 77% in the control arm, leading to a pooled estimate of 78.5%. The antibiotic rates for the sample size re-estimation are estimated in a blinded fashion as described in Kieser et al. [23] as 86% in the experimental arm and 71% in the control arm. In case of a clinical failure rate of 2.5%, the blinded re-estimated power for the co-primary endpoint is around 55% for 1365 patients per arm.

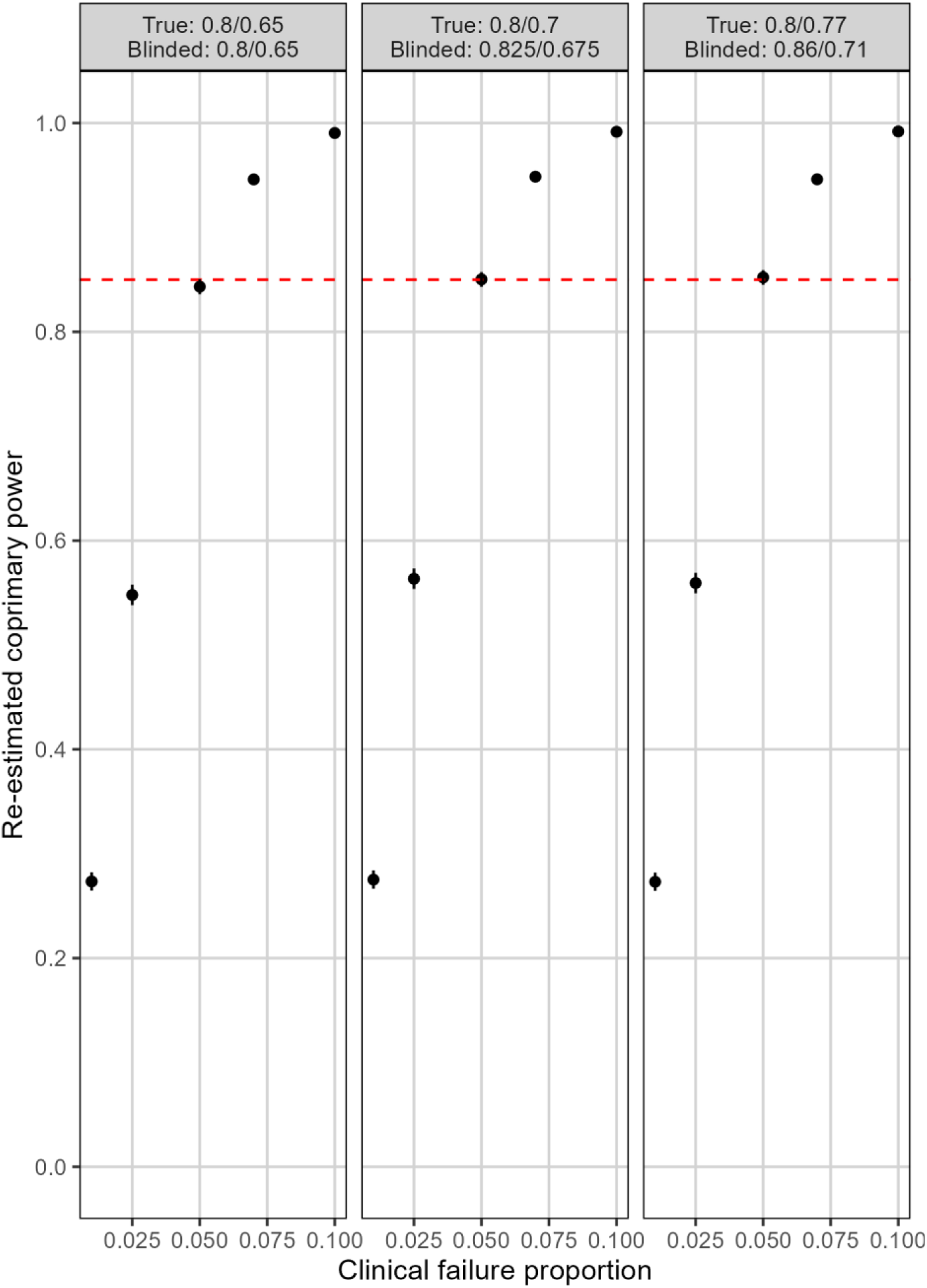

### 10.2 Type I error results

Because the non-inferiorty margin is a relative function of the clinical failure rate we assessed the type-I error as discussed in Quartagno et al. [24]. We conclude that the type-I error was not inflated.

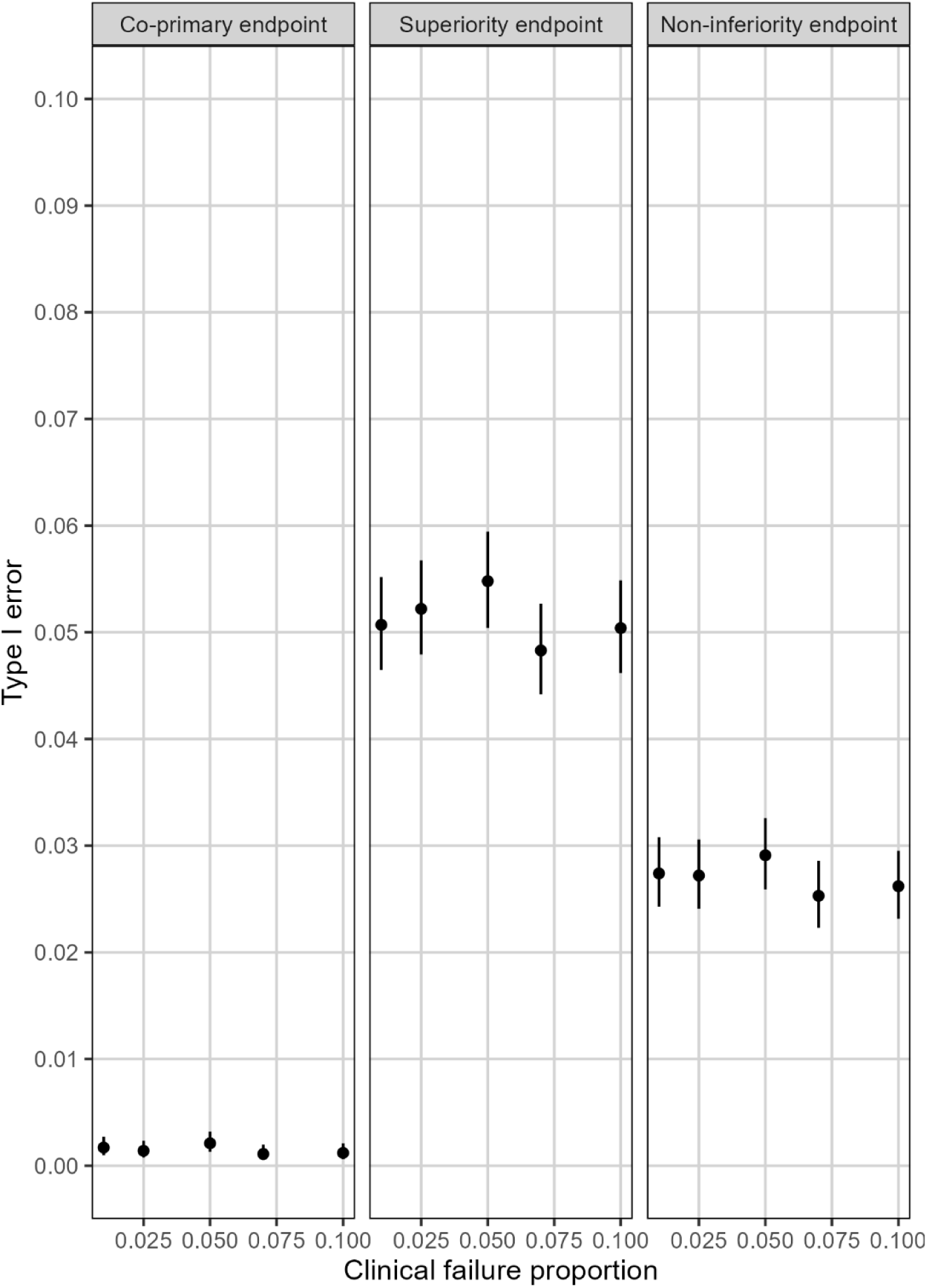

### 10.3 Early stopping for futility: power

Because early stopping for futility affects the power of the co-primary endpoint, we assessed the power under futility stopping if the conditional power at the interim analysis is lower than 20%.

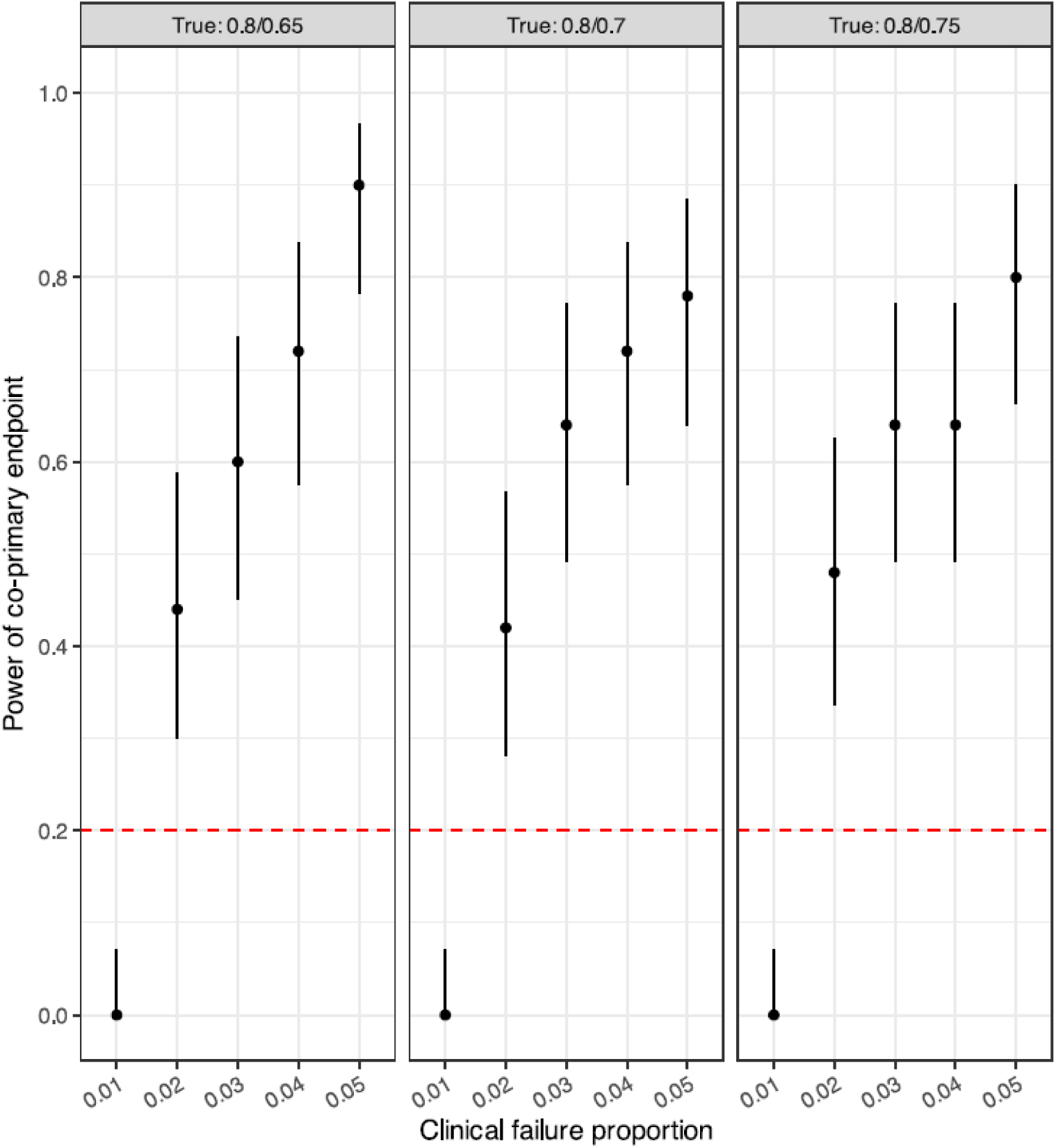

